# Characterizing the Dynamic of COVID-19 with a New Epidemic Model: Susceptible-Exposed-Symptomatic-Asymptomatic-Active-Removed

**DOI:** 10.1101/2020.12.08.20246264

**Authors:** Grace Y Yi, Pingbo Hu, Wenqing He

## Abstract

The coronavirus disease 2019 (COVID-19), caused by the severe acute respiratory syndrome coronavirus 2 (SARS-CoV-2), has spread stealthily and presented a tremendous threat to the public. It is important to investigate the transmission dynamic of COVID-19 to help understand the impact of the disease on public health and economy. While a number of epidemic models have been available to study infectious diseases, they are inadequate to describe the dynamic of COVID-19. In this paper, we develop a new epidemic model which utilizes a set of ordinary differential equations with unknown parameters to delineate the transmission process of COVID-19. Different from the traditional epidemic models, this model accounts for asymptomatic infections as well the lag between symptoms onset and the confirmation date of infection. We describe an estimation procedure for the unknown parameters in the proposed model by adapting the *iterated filter-ensemble adjustment Kalman filter* (IF-EAKF) algorithm to the reported number of confirmed cases. To assess the performance of our proposed model, we examine COVID-19 data in Quebec for the period of April 2, 2020 to May 10, 2020 and carry out sensitivity studies under a variety of assumptions. To reflect the transmission potential of an infected case, we derive the *basic reproduction number* from the proposed model. The estimated basic reproduction number suggests that the pandemic situation in Quebec for the period of April 2, 2020 to May 10, 2020 is not under control.

## 1 Introduction

The coronavirus disease 2019 (COVID-19), caused by the severe acute respiratory syndrome coronavirus 2 (SARS-CoV-2), has spread stealthily and has present a tremendous threat to the public health. On March 11, 2020, the World Health Organization (WHO) declared COVID-19 to be a global pandemic. The first COVID-19 case in Canada was identified in Toronto on January 25, 2020 (The Canadian Press, 2020). In the early stage, imported cases from other countries were the main source of the COVID-19 outbreak in Canada. On March 16, 2020, the closure of Canadian borders was announced to people who are not Canadian citizens or permanent residents (Vogel, 2020). Various measures and interventions have been taken by the federal government and provincial governments to mitigate the virus spread.

While those steps are important to help contain the virus transmissions, it is imperative to investigate the transmission dynamic from the quantitative perspectives. In the literature, a variety of epidemic models have been developed to study infectious diseases, including the *Susceptible-Infectious-Recovered* (SIR) model (e.g., Kermack and McKendrick, 1927), the *Susceptible-Infectious-Susceptible* (SIS) model (e.g., Duan et al., 2015), the *Susceptible-Exposed-Infectious-Recovered* (SEIR) model (e.g., Duan et al., 2015), the Reed-Frost model (e.g., Abbey, 1952), and their variants (e.g., Ng and Orav, 1990; Ng, Turinici, and Danchin, 2003). Applications of those epidemic models have been extensive. To name a few, Osthus et al. (2017) used the SIR model to forecast seasonal influenza; Shah and Gupta (2013) applied the SEIR model to examine transmission processes of vector borne diseases; Ng and Orav (1990) proposed a generalized Reed-Frost model to predict human immunodeficiency virus (HIV) incidence in San Francisco’s homosexual population; and Ng et al. (2003) developed a modified SEIR model, called the *Susceptible-Exposed-Infectious-Recovered-Protection* (SEIRP) model, to study the outbreak of *Severe Acute Respiratory Syndrome* (SARS) in China.

While many epidemic models are available, they are inadequate to facilitate the unique epidemiological characteristics of COVID-19. In this paper, we propose a new epidemic model, called the *Susceptible-Exposed-Asymptomatic-Symptomatic-Active-Removed* (SEASAR) model, to delineate the COVID-19 transmission dynamic. This model generalizes the SIR and SEIR models by accounting for asymptomatic infections and the lag between symptoms onset and the diagnosis time, yet it does not require more assumptions required by the SIR and SEIR models. Consistent with many epidemic models, we make two rountine assumptions: (1) the population is homogeneous and well-mixed; and (2) there are no inbound and outbound travels. Similar to the SIR and SEIR models, the SEASAR model is a deterministic model which utilizes differential equations to describe the transmission dynamic of COVID-19. Furthermore, we derive the basic reproduction number from the proposed model to provide a scalar measure of the pandemic.

To implement the proposed model, we develop an estimation procedure for the model parameters by adapting the iterated filter-ensemble adjustment Kalman filter (IF-EAKF) algorithm (e.g., Li et al., 2020), where resampling from Bayesian posterior distributions is basically invoked. We evaluate the performance of our transmission model and the estimation algorithm by analyzing the COVID-19 data in Quebec for the period of April 2, 2020 to May 10, 2020, where we focus on comparing differences between the predicted cumulative numbers of cases and the reported cumulative numbers of cases. We also conduct sensitivity analyses to assess how the estimation of the model parameters and the predicted results may change as the model assumptions are altered.

Analyzing the COVID-19 data in Quebec is driven by the following considerations. Quebec is the worst-hit province in Canada, and it is thereby interesting to examine the virus transmissions in this province. More importantly, it is imperative to ensure the required conditions to be met as much as possible when applying the model to analyze data. As pointed out earlier, the validity of the proposed SEASAR model hinges on the assumption of no inbound and outbound travels, which is typically untrue in practice, but modeling data in a certain period is likely to make this assumption approximately true. For the period of April 2, 2020 to May 10, 2020, Quebec government set up checkpoints to block all non-essential travels into the province and people in Quebec were advised to stay home, thus, inbound and outbound travels in Quebec for this time window are perceived to be the least.

The remainder of this article is organized as follows. We develop the SEASAR model and elaborate on its rationale in Section 2. In Section 3, we present the initialization setup of the SEASAR model and describe the implementation procedure by adapting the IF-EAKF algorithm. In Section 4, we utilize the proposed SEASAR model to analyze the Quebec COVID-19 data for the period of April 2, 2020 to May 10, 2020. The article is concluded with a discussion presented in Section 5.

## 2 Model Framework

With a meta analysis, He et al. (2020) estimated that about 46% individuals with COVID-19 are asymptomatic, and they have the infectious ability to transmit the disease (e.g., Hao et al., 2020; Li et al., 2020). Due to the limited testing resources and the flu-like manifestation of COVID-19 as well as the incubation period, there is a time lag between symptoms onset and being confirmed for infected individuals (Kramer et al., 2020). To facilitate these features of COVID-19, we develop a new model, called the *Susceptible-Exposed-Asymptomatic-Symptomatic-Active-Removed* (SEASAR) model, under the assumptions of a well-mixed homogeneous population and of no inbound and outbound travels (i.e., the population size remains to be unchanged over time).

### 2.1 Illustration of the Proposed Model

To illustrate the ideas, we first consider a *static* framework by focusing at a given time point, say, on a given day. We divide the target population into six subpopulations with specific features, where *S* represents the subpopulation of *susceptible* cases (i.e., those at risk of becoming infected with the novel coronavirus), *E* is the subpopulation of *exposed* cases (i.e., those who are infected but do not have the infectious ability yet and are still in the latent period (e.g., Peng et al., 2020), *I*_*a*_ stands for the subpopulation of *asymptomatic* infections (i.e., those with the infectious ability but showing no symptoms), *I*_*s*_ represents the subpopulation of *symptomatic* infections (i.e., those who show symptoms and have the infectious ability but are not confirmed yet), *A* is the subpopulation of *active* cases (i.e., those confirmed cases who do not recover or die), and *R* denotes the subpopulation of *removed* cases (i.e., those confirmed cases who recover or die from COVID-19).

Next, we introduce the parameters to facilitate the dynamic changes among the subpopulations. Let *Z* denote the *average latent period*, defined as the average time from being infected to having the infectious ability. Various studies have been conducted to estimate the value of *Z* (e.g., Bai et al., 2020; Guan et al., 2020; He, Yi, and Zhu, 2020), so here we take *Z* as being available. Let *θ* denote the *symptomatic transmission rate*, defined as the average number of individuals infected by a symptomatic case per unit time (e.g., a day). Let the *asymptomatic transmission rate* be denoted as *µθ*, which is defined as the average number of individuals infected by an asymptomatic case per unit time. As asymptomatic infections are regarded as less infectious than symptomatic cases (e.g., Li et al., 2020), *µ* is a constant between 0 and 1. Let *α* denote the fraction of *symptomatic* infections relative to all infections, let *β* denote the average rate for *asymptomatic* infections to develop symptoms per unit time, and let *γ* denote the average recovery rate of *asymptomatic* infections per unit time. Let *F* stand for the average time from symptoms onset to the time of being confirmed, let *B* denote the average time from being confirmed to being recovered, and let *J* represent the average time from being confirmed to die. Figure 1 is a flow chart showing the relationship among the six subpopulations. A black solid arrow between two subpopulations indicates the members of one subpopulation can move into the other subpopulation; a red dashed arrow between two subpopulations indicates that members in one subpopulation can be infected by members in the other subpopulation. Once COVID-19 cases are confirmed, they will be quarantined and lose the infectious ability, so the group *S* cannot be infected by the group *A*. Due to the limited test capacity, asymptomatic individuals are not being tested for COVID-19 (especially in the early stage of the outbreak). Thus, there is no transition from *I*_*a*_ to *A*. The parameters on black solid lines determine the number of people who change from one state to another state per unit time; and the parameters on red dashed lines determine the number of people infected by asymptomatic or symptomatic cases per unit time.

**Figure 1:**
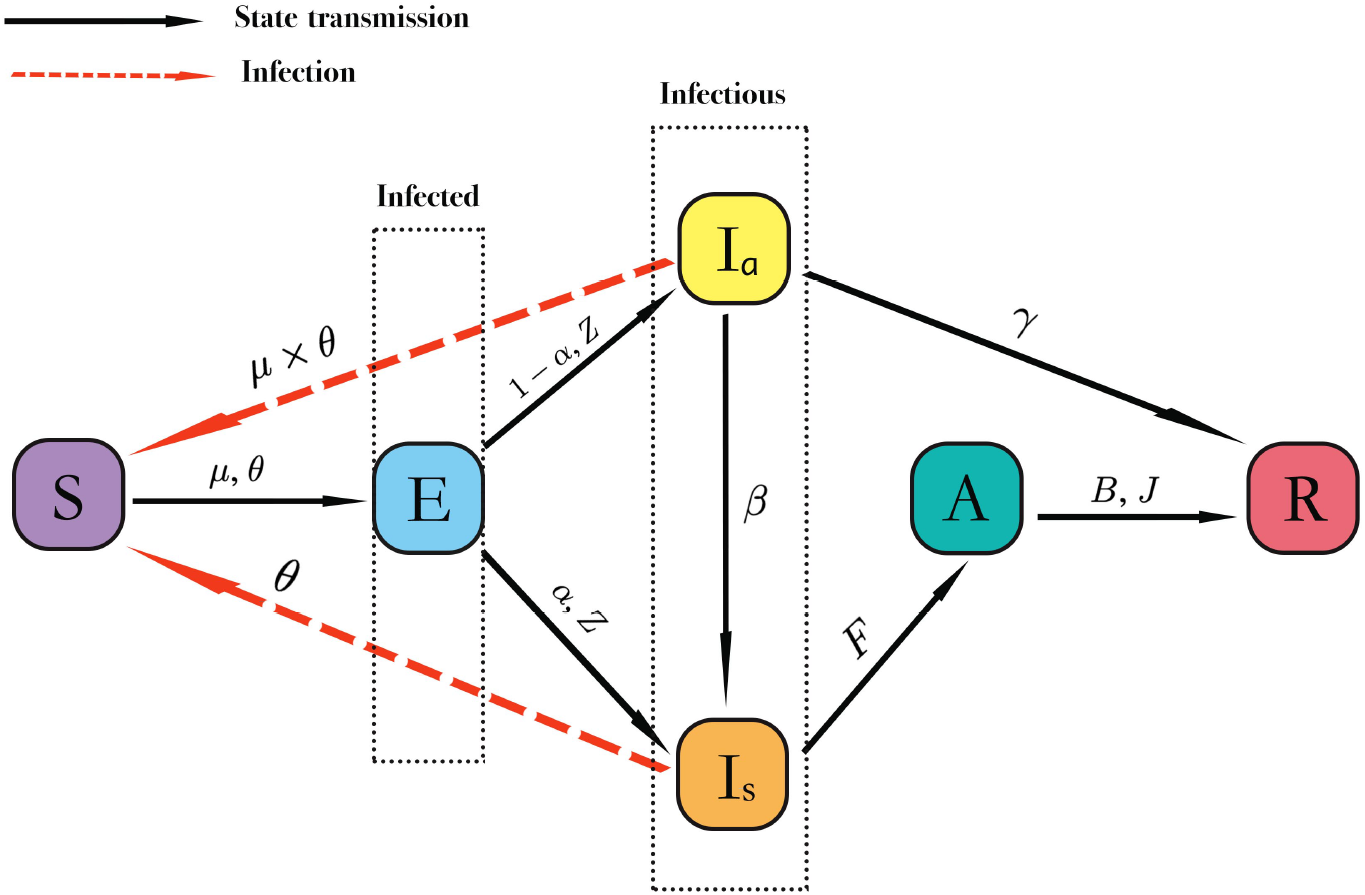
Illustration of the SEASAR model. The population is divided into six compartments: S (susceptible), E (exposed), I_a_ (asymptomatic), I_s_ (symptomatic), A (active), and R (removed).

### 2.2 Dynamic Model

Figure 1 shows a static chart for the transmissions among the six subpopulations at a given time point. However, for any time period, the transmissions are not static but dynamic. To characterize the *dynamic* evolution of the subpopulations over time, we modify the six subpopulations by showing their dependence on time *t*, yielding the state components {*S*(*t*), *E*(*t*), *I*_*a*_(*t*), *I*_*s*_(*t*), *A*(*t*), *R*(*t*)}. For ease of notation, we also use the same symbols to represent the *sizes* of those state components. Let *ϕ*(*t*) = (*S*(*t*), *E*(*t*), *I*_*a*_(*t*), *I*_*s*_(*t*), *A*(*t*), *R*(*t*))^T^ denote the vector of the six subpopulation sizes at time *t*.

Given the setup, we now present the proposed SEASAR model, given by the following set of ordinary differential equations:

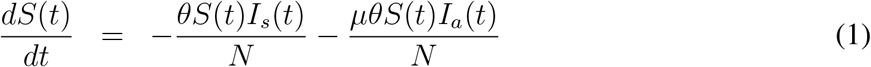

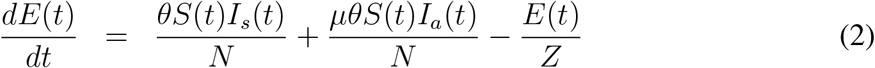

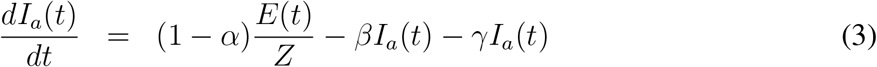

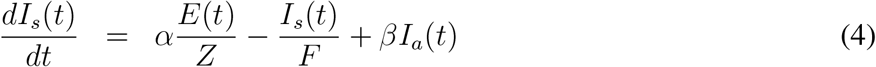

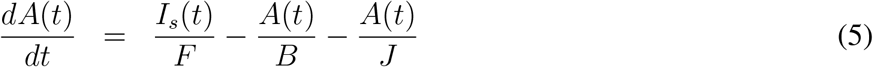

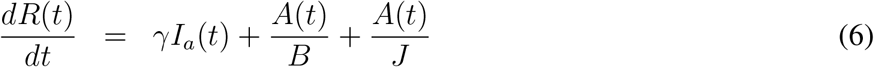

where *N* is the population size, which is time-invariant due to the assumptions of no outbound and inbound travels. Because of this assumption, *S*(*t*)+*E*(*t*)+*I*_*a*_(*t*)+*I*_*s*_(*t*)+*A*(*t*)+*R*(*t*) = *N* for any time point *t*, and hence, any equation above is determined by other five equations.

For ease of referral of equations (1)-(6) in the later development, we express those equations in a compact form:

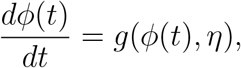

where *g*(·, ·) represents the vector function determined by the right hand side of (1)-(6). Figure 2 is a flowchart of the transmission dynamic for the six subpopulations together with the associated values.

**Figure 2:**
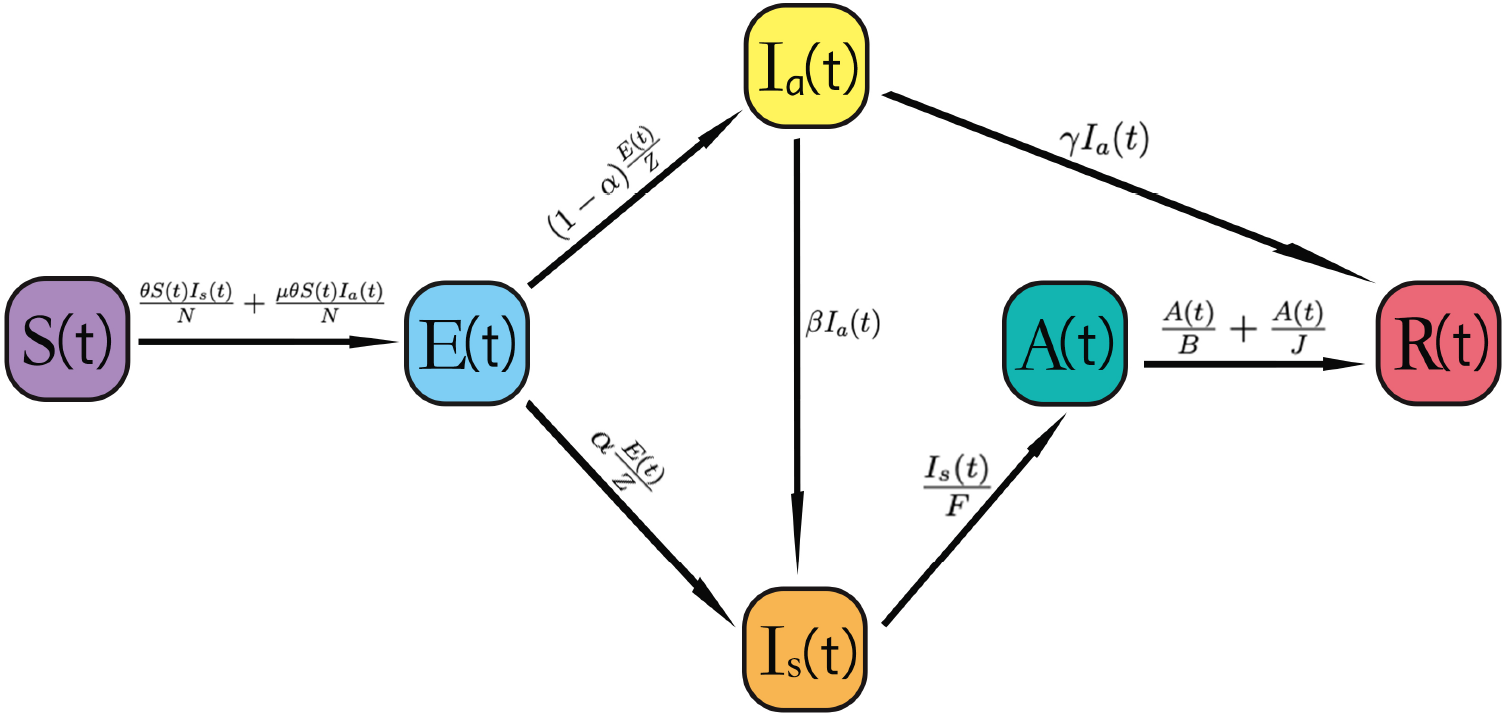
Dynamic of the transmission among the six subpopulations at any time t

The right hand side of each of equations (1)-(6) shows how the change rate of each subpopulation is related to the transmission rates as well as the associated parameters defined earlier. To see why, we first examine the right hand side of (1) by considering a small time interval [*t, t* + Δ*t*) of length Δ*t*. Over the time interval [*t, t* + Δ*t*), because *θI*_*s*_(*t*)Δ*t* represents the average number of people infected by symptomatic infections and 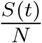 is the proportion of susceptible cases, so 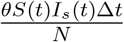 equals the average number of *susceptible* cases who are infected by *symptomatic* infections; similarly, 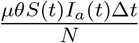 equals the average number of *susceptible* cases who are infected by *asymptomatic* infections. Thus, 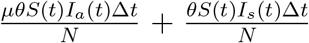 records the average number of *susceptible* cases moving to the *exposed* state over the time interval [*t, t* + Δ*t*), i.e., the reduced number of *susceptible* cases over the time interval [*t, t* + Δ*t*) is 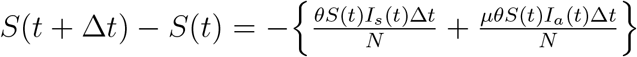. Then dividing Δ*t* on the both sides and letting Δ*t* → 0 yields (1).

Equations (2)-(6) can be illustrated in an analogous way. Regarding (2), due to the changing from susceptible cases to being in the *exposed* state, the number of *exposed* cases is increased by 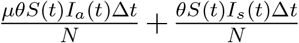 over the interval [*t, t* + Δ*t*); yet due to the latent period, the number of *exposed* cases is reduced by 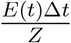 over the interval [*t, t* + Δ*t*), yielding that the difference in the number of *exposed* cases over the time interval [*t, t* + Δ*t*) is 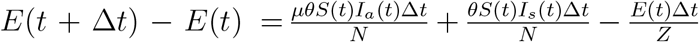. Dividing Δ*t* on the both sides and letting Δ*t* → 0 yields (2).

Regarding (3), when *exposed* cases pass the latent period, they become either *asymptomatic* or *symptomatic* infections. As the fraction of *asymptomatic* infections is 1 − *α*, the number of asymptomatic infections is thus increased by 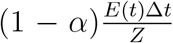 over the interval [*t, t* + Δ*t*). On the other hand, *βI*_*a*_(*t*)Δ*t* is the average number of infections who initially are *asymptomatic* but later show symptoms over the interval [*t, t* + Δ*t*) (i.e., people in this group turn from the *asymptomatic* state to the *symptomatic* state), and *γI*_*a*_(*t*)Δ*t* is the average number of *asymptomatic* infections who recover from COVID-19 (i.e., people in this group move from the *asymptomatic* state to the *removed* state). Thus, the number of asymptomatic infections is reduced by *βI*_*a*_(*t*)Δ*t* + *γI*_*a*_(*t*)Δ*t* over the interval [*t, t* + Δ*t*). Therefore, the difference in the number of asymptomatic infections is 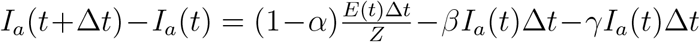, yielding (3) by dividing Δ*t* on the both sides and letting Δ*t* → 0.

Regarding (4), due to the change of individuals from the *exposed* and *asymptomatic* states to the *symptomatic* state, the number of symptomatic infections is increased by 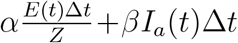 over the interval [*t, t* + Δ*t*). On the other hand, 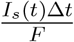 is the average number of *symptomatic* infections who are confirmed over the interval [*t, t* + Δ*t*) (i.e., people in this group turn from the *symptomatic* state to the *active* state); in other words, the reduced number of *symptomatic* infections over the interval [*t, t* + Δ*t*) is 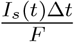. Thus, the difference in the number of *symptomatic* infections is 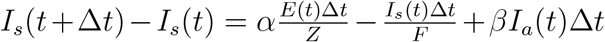, leading to (4) if dividing Δ*t* on the both sides and letting Δ*t* → 0.

Regarding (5), due to the changing from the *symptomatic* state to the *active* state, the number of active cases is increased by 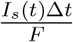 over the interval [*t, t* + Δ*t*). Since 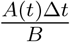 is the average number of *active* cases who recover from COVID-19 and 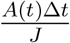 is the average number of *active* cases who die from COVID-19 over the interval [*t, t* + Δ*t*) (i.e., people in the two groups turn from the *active* state to the *removed* state), the number of active cases is reduced by 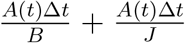 over the interval [*t, t* + Δ*t*). Therefore, the difference in the number of active cases is 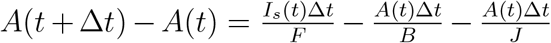, yielding (5) if we divide Δ*t* on the both sides and let Δ*t* → 0.

Regarding (6), due to the changing from the *active* and *asymptomatic* state to the *removed* state, the number of removed cases is increased by 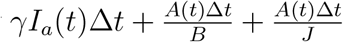 over the interval [*t, t* + Δ*t*), i.e., 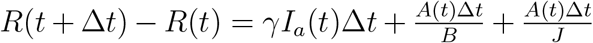. Thus, dividing Δ*t* on the both sides and letting Δ*t* → 0 yields (6). Alternatively, (6) equals the negative sum of (1) to (5).

### 2.3 Basic Reproduction Number

Let *η* = (*θ, µ, α, β, γ, F, B, J*)^T^ denote the vector of parameters of prime interest. Knowing the value of *η* allows us to describe the six subpopulations sizes using the models (1)-(6). Further, it enables us to describe the severity of the pandemic using some simple measure such as the *basic reproduction number*, denoted *R*_0_, which is defined as the expected number of cases infected by one case in a population consisting of individuals susceptible to infection.

A large value of *R*_0_ indicates a more severe pandemic. Usually, comparing *R*_0_ to 1 describes the spread of the disease. “*R*_0_ > 1” suggests that the infection is spreading in the population, and “*R*_0_ < 1” indicates the dying down situation. In Appendix A, we show that the mathematical expression of *R*_0_ derived from the SEASAR model is

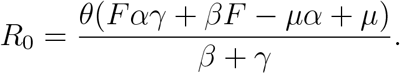

## 3 Estimation Procedure

In this section, we develop an estimation procedure for *η* by adapting the iterated filter-ensemble adjustment Kalman filter (IF-EAKF) algorithm (e.g., Li et al., 2020) in combination with the 4th order Runge-Kutta (RK4) method (e.g., Süli and Mayers, 2003, p.328).

### 3.1 Initial Sizes of Subpopulations

Let *τ*_0_ denote the initial time point from which we start examining the data. By time *τ*_0_, let *R*_*c*_, *C*_0_ and *D*_0_ denote the reported cumulative number of recoveries, confirmed cases and deaths from COVID-19, respectively, which are available; and let *R*_*a*_ denote the cumulative number of recovered asymptomatic cases, which is unavailable. Let *Q*(*t*) denote the number of reported cases with symptoms onset on day *t*.

To facilitate the relationship between the observed and unobserved values, let 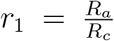 denote the ratio of the unobserved *R*_*a*_ to the observed *R*_*c*_, and let 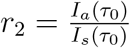 represent the ratio of unobserved size *I*_*a*_(*τ*_0_) to the reported size *I*_*s*_(*τ*_0_). Motived by Hao et al. (2020), we express the size *E*(*τ*_0_) in terms of its relative value to the total number of reported cases with symptoms onset over the time window of length 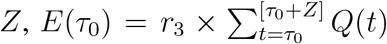, where the function [*x*] represents the biggest integer that is less than or equal to *x*, and *r*_3_ is a positive value.

While the introduction of the ratios *r*_1_, *r*_2_ and *r*_3_ does not give us a way to determine the unobserved values *R*_*a*_, *I*_*a*_(*τ*_0_) and *E*(*τ*_0_) using the observed data, these ratios offer us convenient measures to describe the pandemic situation in relative scales of the observed values at time *τ*_0_. For instance, at the early stage of the pandemic, the testing kits are limited, so the number of recoveries from *confirmed* cases is likely to be a lot smaller than that from *asymptomatic* individuals, suggesting a large value of *r*_1_. If *r*_2_ is bigger than 1, then there are more *asymptomatic* infections than *symptomatic* infections.

Table 1 summarizes the initial sizes of the six subpopulations which are to be used in Section 3.3, where the values of *r*_1_, *r*_2_ and *r*_3_ are specified. Write *ϕ*(*τ*_0_) = (*S*(*τ*_0_), *E*(*τ*_0_), *I*_*a*_(*τ*_0_), *I*_*s*_(*τ*_0_), *A*(*τ*_0_), *R*(*τ*_0_))^T^.

**Table 1:** Initial state size for the SEASAR model

| Variable | Meaning | Value |
| --- | --- | --- |
| $S(\tau_0)$ | The initial number of susceptible cases | $N - E(\tau_0) - I_a(\tau_0) - I_s(\tau_0) - A(\tau_0) - R(\tau_0)$ |
| $E(\tau_0)$ | The initial number of exposed cases | $r_3 \times \sum_{t=\tau_0}^{\lceil \tau_0 + Z \rceil} Q(t)$ |
| $I_a(\tau_0)$ | The initial number of asymptomatic infections | $r_2 \{ \sum_{t \leq \tau_0} Q(t) - C_0 \}$ |
| $I_s(\tau_0)$ | The initial number of symptomatic infections | $\sum_{t \leq \tau_0} Q(t) - C_0$ |
| $A(\tau_0)$ | The initial number of active cases | $C_0 - R_c - D_0$ |
| $R(\tau_0)$ | The initial number of removed cases | $D_0 + (1 + r_1)R_c$ |

### 3.2 Initialization of Model Parameters

Estimation of *η*, to be described in Section 3.3, is conducted by an iterative approximation procedure using the posterior distributions of *η*. Here we assume the prior information for the parameters to be non-informative except for constraining the parameters to certain ranges to reflect our a priori knowledge about them.

To be specific, the transmission rate *θ* of symptomatic infections is considered to be 0 ≤ *θ* ≤ 7 to cover the reported values in the literature, including Hao et al. (2020) and Li et al. (2020). The multiplicative factor *µ* is assumed to satisfy 0.1 ≤ *µ* ≤ 1, the fraction *α* of symptomatic infections relative to all infections is restricted to be 0.1 ≤ *α* ≤ 1, the average rate *β* of asymptomatic infections who develop symptoms per unit time is constrained to be 0.0002 ≤ *β* ≤ 0.8, and the average recovery rate *γ* of asymptomatic infections per unit time is set as 0.1 ≤ *γ* ≤ 1. The average time *F* from symptoms onset to being confirmed is considered to be between 1 and 10 days based on the study of Kramer et al. (2020). Based on the WHO report (WHO, 2020), the average time *B* from being confirmed to being recovered is considered to be in the range of 7 to 42 days, and the average time *J* from being confirmed to die is taken to change from 14 to 56 days.

### 3.3 IF-EAKF algorithm

With the setup in Sections 3.1 and 3.2, we describe an estimation procedure by adapting the iterated filtering (IF) algorithm. The IF approach basically produces the maximum likelihood estimates of model parameters and has been successfully applied to study many infectious diseases (Li et al., 2020), including cholera (e.g., King et al., 2008) and measles (e.g., He et al., 2010). An efficient IF approach roots in using the *ensemble adjustment Kalman filter* (EAKF) which can generate satisfactory results with only hundreds of samples (Li et al., 2020).

Since the number of confirmed cases is reported on a daily basis, we take the time unit as a day. Consider a population of interest. In contrast to the number *Q*(*t*) of reported cases with symptoms onset on day *t*, defined in Section 3.1, we let *Y* (*t*) denote the number of confirmed cases to be reported on day *t*, which is regarded as a random variable, and let *y*_*t*_ denote its realization. Let 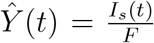 denote the number of confirmed cases on day *t* that is generated from the SEASAR model. We assume that *Y* (*t*) and *Ŷ*(*t*) are connected through the model

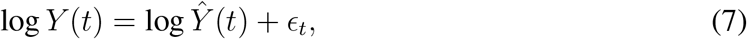

where the *ϵ*_*t*_ are independent of each other and of the *Ŷ*(*t*), *ϕ*(*t*) and *η*; and *ϵ*_*t*_ follows a Gaussian distribution with mean 0 and time-dependent variance 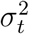.

Being called the *observational error variance* (OEV) by Li et al. (2020), 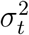 is often estimated heuristically via an assumed function form of the observations. For example, in the analysis in Section 4.2, 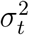 is assumed to be

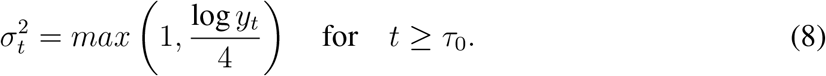

Similar forms of OEV were used in the studies of other infectious diseases, including influenza (e.g., Pei et al., 2018), Ebola (e.g., Shaman et al., 2014), West Nile virus (e.g., DeFelice et al., 2017), and respiratory syncytial virus (e.g., Reis and Shaman, 2016).

The model parameter *η* for (1)-(6), the subpopulation sizes *ϕ*(*t*) and *Ŷ*(*t*) at time *t* for *t* ≥ *τ*_0_ are unknown. One may consider the joint posterior distribution of *η*, log *Ŷ*(*t*) and log *ϕ*(*t*), and then use the mean of the posterior distribution of *η* to estimate *η*, where log *ϕ*(*t*) := (log *S*(*t*), log *E*(*t*), log *I*_*a*_(*t*), log *I*_*s*_(*t*), log *A*(*t*), log *R*(*t*))^T^. To reduce computation costs, we adapt the discussion of (Anderson, 2001, p.2888) and describe a simple iterative procedure by pairing two members in {*η*, log *Ŷ*(*t*), log *ϕ*(*t*)}, where we typically pair log *Ŷ*(*t*) with each component in {*η*, log *ϕ*(*t*)}. Let *τ*_0_ + *T* − 1 denote the time point by which we stop iterations. To see the main ideas, here we elaborate on the detail of the first iteration for the IF-EAKF algorithm which includes the following six stages, with similar details for other iterations omitted.

- **Stage 1:** At time *t* = *τ*_0_, generate prior values for *η* and *Ŷ*(*t*):
  − **Step 1:** Let *π*_*η*_ denote a prior distribution for parameter *η*, which is taken as the uniform distribution over the ranges specified in Section 3.2, with the assumption that the parameter components in *η* are independent of each other.
  − **Step 2:** Specify a positive integer, say *n*. Then generate *n* values from *π*_*η*_, denoted 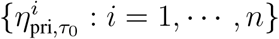, where for each 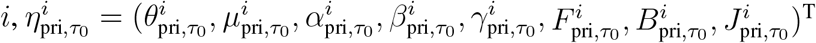.
  − **Step 3:** Using the initial size *I*_*s*_(*τ*_0_) of symptomatic infections, we generate *n* prior values for *Ŷ*(*τ*_0_), denoted 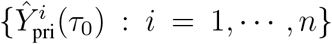, by setting 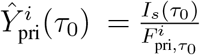 for *i* = 1, …, *n*. Then calculate the sample mean and variance:

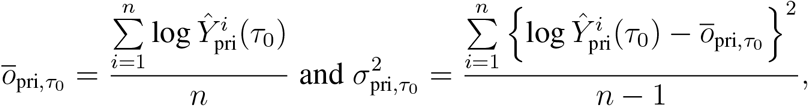

together with the pairwise sample covariance

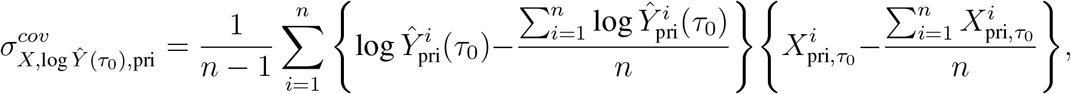

where *X* is a symbol for *θ, µ, α, β, γ, F*, *B*, and *J*.
- **Stage 2:** At time *t* = *τ*_0_, generate posterior values for *η* and *Ŷ*(*t*): We employ the following steps to generate *n* posterior values for *Ŷ*(*τ*_0_) and *η* from their posterior distribution, derived in Appendix B.
  − **Step 1:** Generate *n* posterior values for *Ŷ*(*τ*_0_), denoted 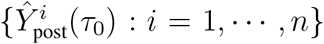 For each 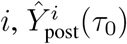 is determined by

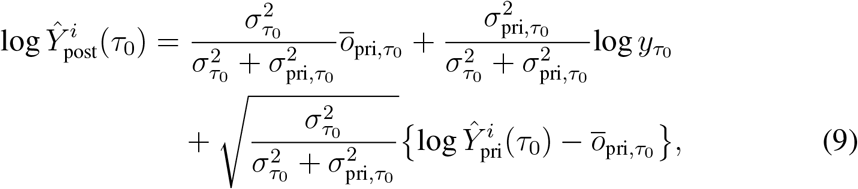

where 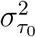 is given by (8) with *t* = *τ*_0_, and 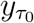 is the number of confirmed cases reported on day *τ*_0_.
  − **Step 2:** Generate *n* posterior values for *η*, denoted 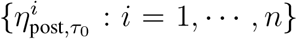, where for each 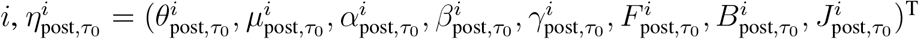 Specifically, each component of 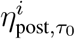 is given by

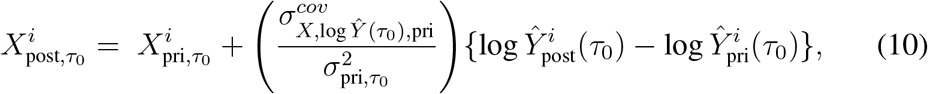

where *X* is a symbol for *θ, µ, α, β, γ, F*, *B*, and *J*.
- **Stage 3:** At time *t* = *τ*_0_ + 1, generate prior values for *η, ϕ*(*t*), and *Ŷ*(*t*):
  − **Step 1:** Generate *n* prior values for *η*, denoted 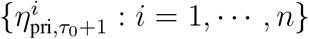, by setting 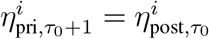 for *i* = 1, …, *n*, where we denote 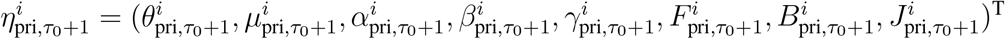, for each *i*.
  − **Step 2:** Using the RK4 method, we generate *n* prior values for *ϕ*(*τ*_0_ + 1), denoted 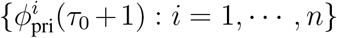, where for each 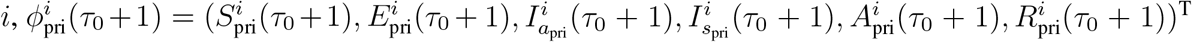. Specifically, let 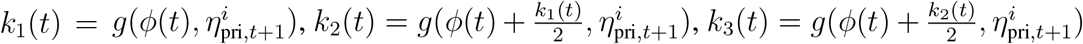, and 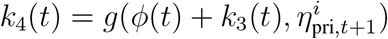, Then we set

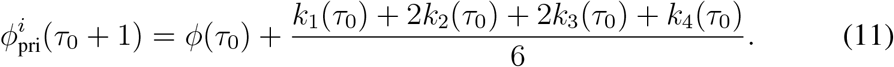
  − **Step 3:** Generate *n* prior values for *Ŷ*(*τ*_0_ +1), denoted 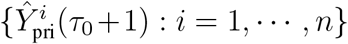, by setting 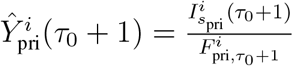 for *i* = 1, …, *n*. Then we calculate

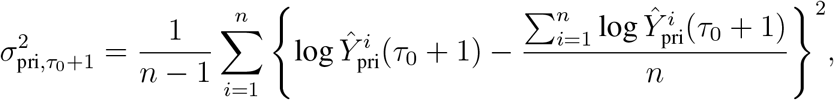

together with the pairwise sample covariance

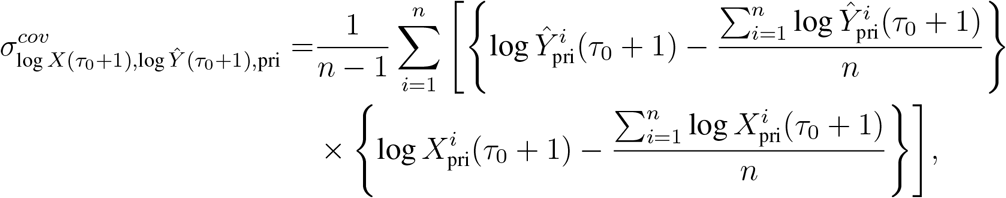

where *X* is a symbol for *S, E, I*_*a*_, *I*_*s*_, *A*, and *R*.
- **Stage 4:** At time *t* = *τ*_0_ + 1, generate posterior values for *η, ϕ*(*t*), and *Ŷ*(*t*). This stage is similar to Stage 2.
  − **Step 1:** Similar to Stage 2, we generate *n* posterior values for *Ŷ*(*τ*_0_ + 1) and *η*, denoted 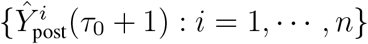 and 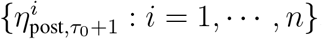, respectively.
  − **Step 2:** Generate *n* posterior values for *ϕ*(*τ*_0_ + 1), denoted 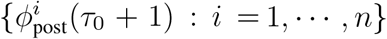, where for each 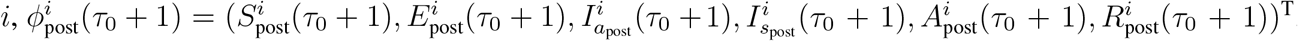. Specifically, each component of 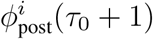 is given by

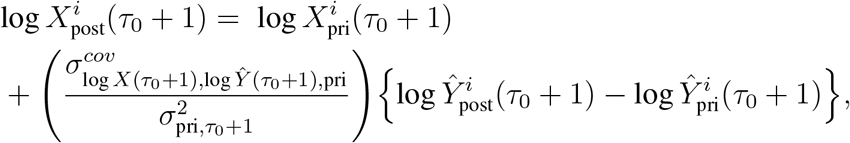

where *X* is a symbol for *S, E, I*_*a*_, *I*_*s*_, *A*, and *R*.
- **Stage 5:** At time *t* = *τ*_0_ + 2, generate prior values for *η, ϕ*(*t*), and *Ŷ*(*t*):
  − **Step 1:** Generate *n* prior values for *η*, denoted 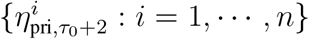, by setting 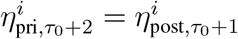 for *i* = 1, …, *n*.
  − **Step 2:** Generate *n* prior values for *ϕ*(*τ*_0_ + 2), denoted 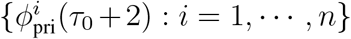, using the RK4 method, where similar to (11),

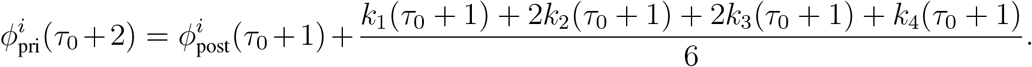
  − **Step 3:** Similar to Step 3 in Stage 3, we generate *n* prior values for *Ŷ*(*τ*_0_ + 2).
- **Stage 6:** Similar to Stage 4, we generate *n* posterior values of *η, ϕ*(*τ*_0_ + 2) and *Ŷ*(*τ*_0_ + 2).

We repeat Stages 5-6 for *t* = *τ*_0_ + 3, …, *τ*_0_ + *T* − 1, and obtain a sequence of posterior values for *η*, denoted 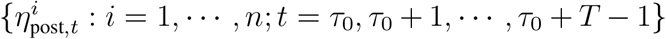. Then we calculate

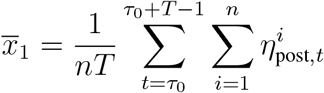

and use *x̄*_1_ for the next iteration.

The proceding descriptions show the steps for the first iteration; steps for subsequent iterations, similar to the first iteration, are outlined in Algorithm 1, where Σ is the covariance matrix of the prior distribution *π*_*η*_ which is shrunk by a discount factor *a* ∈ (0, 1), usually pre-specified to reduce the variance of the posterior distribution of the parameters as the iteration progresses. Using a discount factor is a standard procedure to ensure that 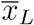 in Algorithm 1, the estimate at the *L*th iteration, is nearly identical to the maximum likelihood estimate of *η*. If *a* is too small, the algorithm may “quench” too fast and fail to find the maximum likelihood estimate; if it is too close to 1, the algorithm may not converge in a reasonable time (see Li et al., 2020, Supplementary Materials, p.8). Practically, *a* can range between 0.9 to 0.99. The number *L* of iterations is set in an ad hoc way, often determined by inspecting the evolution of the posterior parameter distributions (see Li et al., 2020, Supplementary Materials, p.8). The scale of *n* is in hundreds, though in principle, the larger the better.

## 4 Data Analysis

### 4.1 Quebec COVID-19 Data

On April 1, 2020, Quebec provincial government in Canada set up checkpoints to block all non-essential travels into the province and advised people in Quebec to stay home. After May 10, 2020, Quebec and other provinces in Canada gradually reopened the economy, yielding inbound and outbound travels in Quebec. Therefore, it is reasonable to perceive inbound and outbound travels in the period of April 2, 2020 to May 10, 2020 in Quebec to be the least, and the assumption of no inbound and outbound travels required by the proposed SEASAR model is relatively feasible for the Quebec COVID-19 data in this period.

Driven by this, we apply the proposed model to analyze the daily reported number of COVID-19 cases in Quebec, Canada, in this period, available at https://www.canada.ca/en/public-health/services/diseases/2019-novel-coronavirus-infection.html. We take the initial time point *τ*_0_ as April 2, 2020 and split the study period into two parts: the period of April 1 to April 30, 2020, and the period of May 1 to May 10, 2020. The data for the first period are used to estimate the model parameters by applying Algorithm 1, and the data in the second period are used to assess the prediction performance of our proposed model.

#### Algorithm 1: IF-EAKF

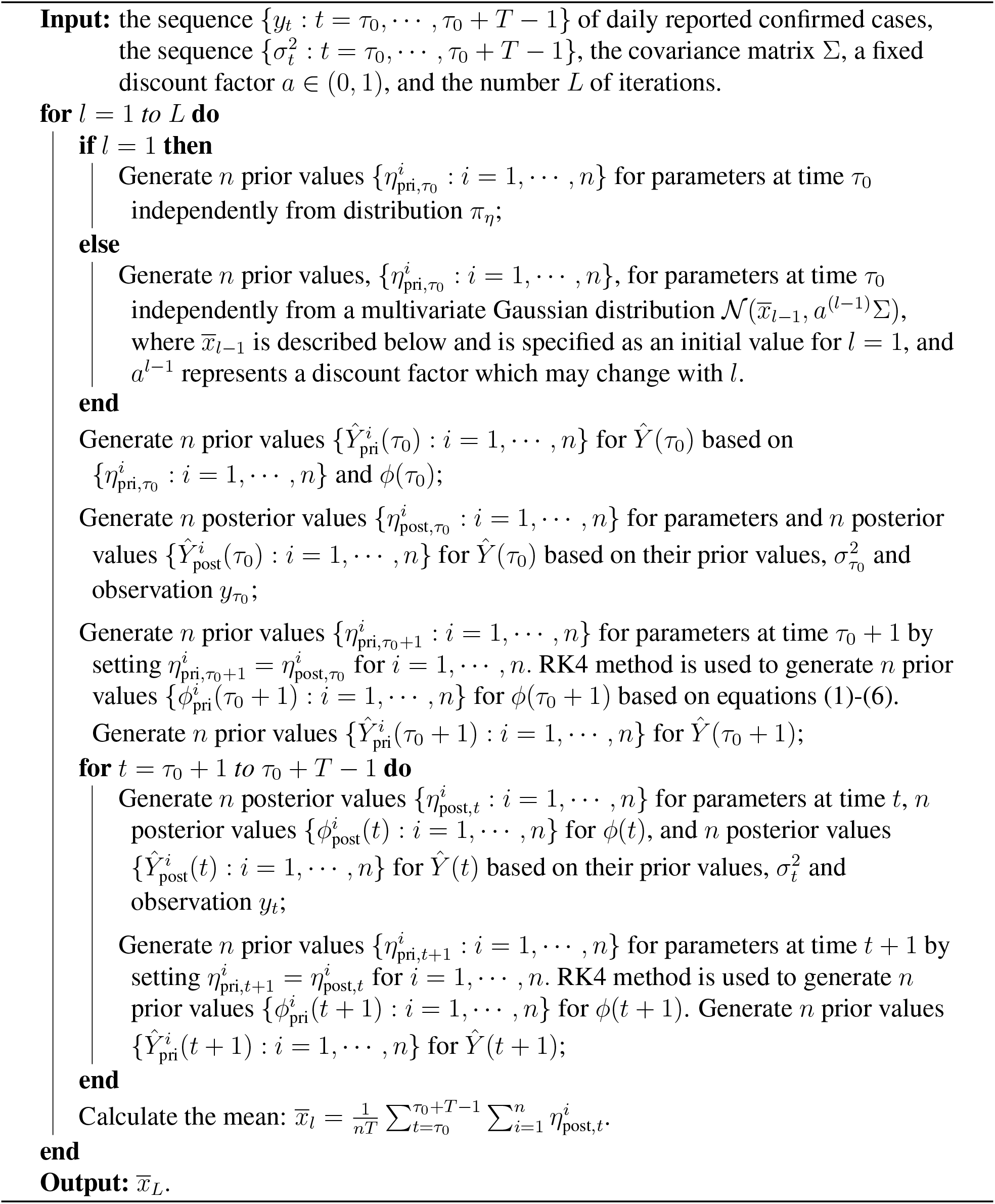

### 4.2 Estimation of Model Parameters

We run Algorithm 1 to the proposed SEASAR model, where we set *n* = 300, *a* = 0.9, *L* = 50, and the average latent period *Z* is taken as 5.2 days, an estimate reported by Bai et al. (2020) and Hao et al. (2020). For the initial sizes of the six subpopulations discussed in Section 3.1, we take *r*_1_ = 2, *r*_2_ = 1, and *r*_3_ = 2. As described in Section 3.1, the initial sizes of subpopulations depend on the daily number of COVID-19 patients with symptoms onset before May 10, 2020, which is not available in Quebec. However, the daily number of COVID-19 patients with symptoms onset in Canada is collected by the Public Health Agency of Canada (https://health-infobase.canada.ca/covid-19/epidemiological-summary-covid-19-cases.html). Using this information, we approximate the daily number of COVID-19 patients with symptoms onset before May 10, 2020 in Quebec. Specifically, let *n*_*t,C*_ denote the number of COVID-19 patients with symptoms onset on day *t* in Canada, and let *m*_*t,C*_ and *m*_*t,Q*_ denote the number of confirmed COVID-19 cases on day *t* in Canada and Quebec, respectively. We then take 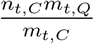 as an estimated number of COVID-19 patients with symptoms onset on day *t* in Quebec.

To account for stochastic effects, we run the IF-EAKF algorithm 1000 times and display in Figure 4 the histograms for those estimates. A 95% confidence interval (CI) for each parameter is determined by using the 2.5% and 97.5% percentiles of the 1000 estimates as its lower and upper bounds, respectively. Table 2 reports the average estimates and the associated 95% CIs for the parameters, together with the average of those 1000 estimated *basic reproduction numbers* and its associated 95% CI. The estimate of *R*_0_ suggests that the pandemic situation in Quebec for the period of April 2, 2020 to April 30, 2020 is not under control and the virus spread continues.

**Figure 3:**
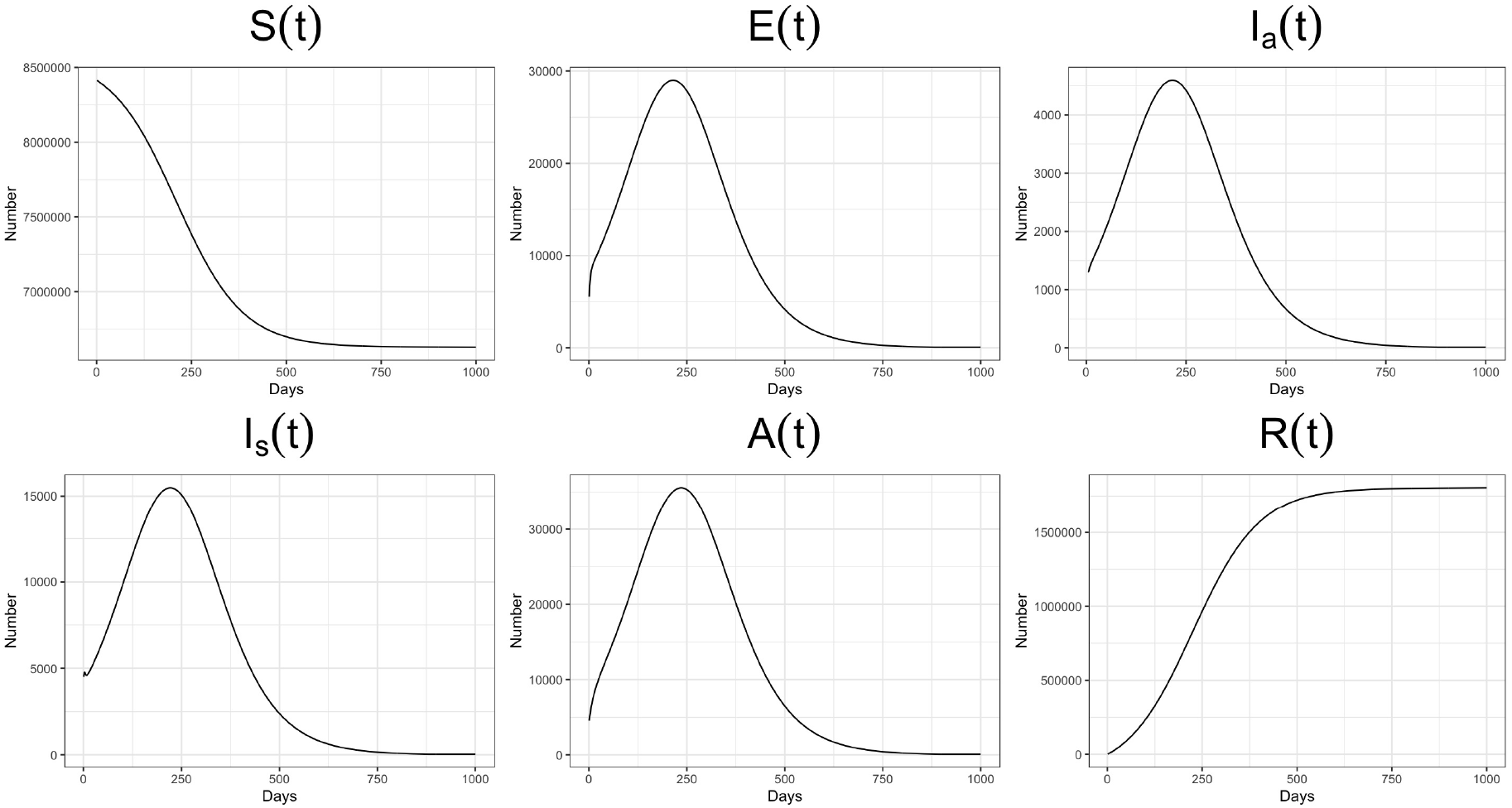
The estimated curves of S(t), E(t), I_a_(t), I_s_(t), A(t), and R(t).

**Figure 4:**
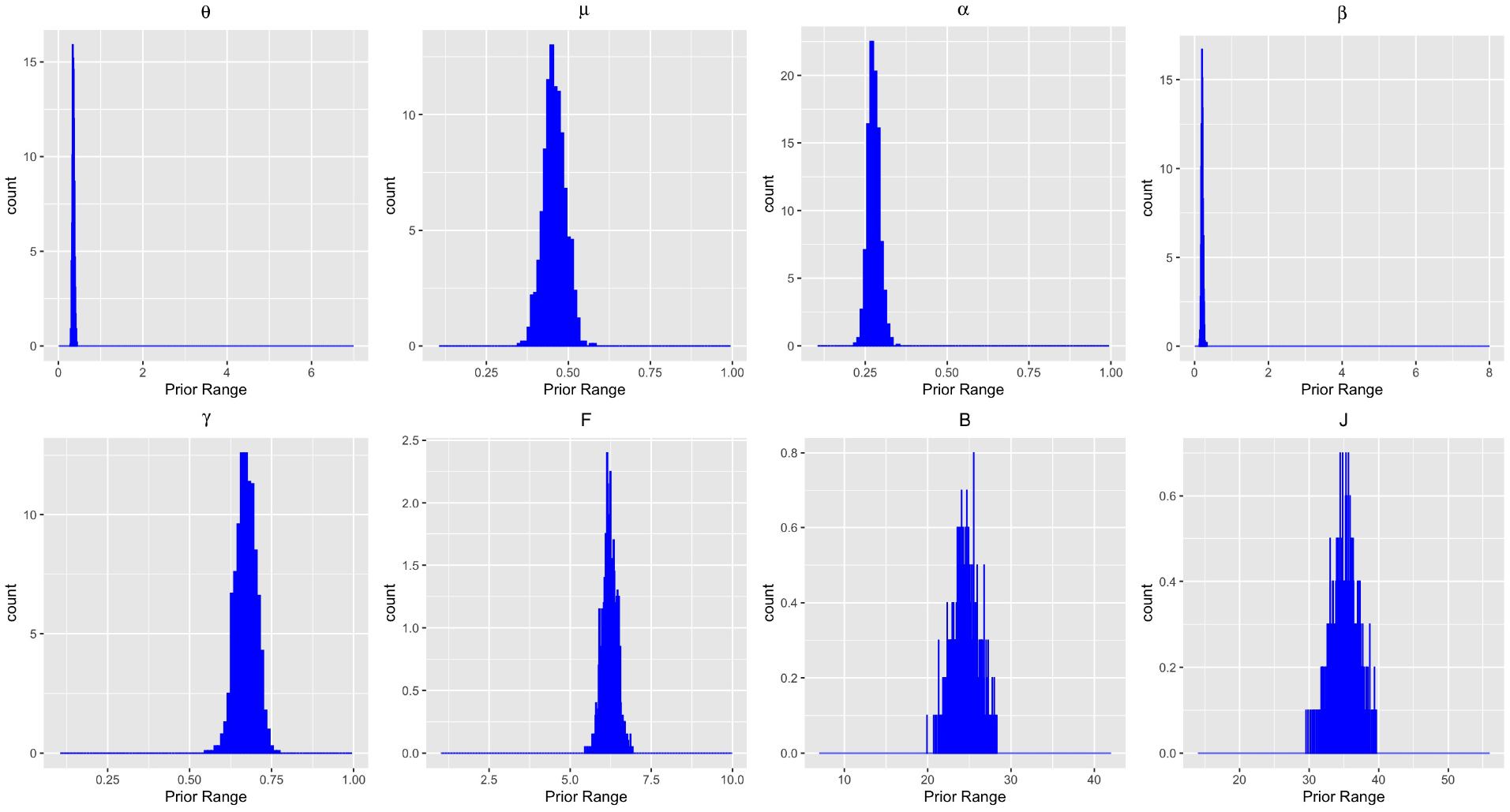
The distribution of 1000 estimates for each parameter. The range of the x-axis for each subfigure is set as the initial range of the corresponding parameter.

**Table 2:**
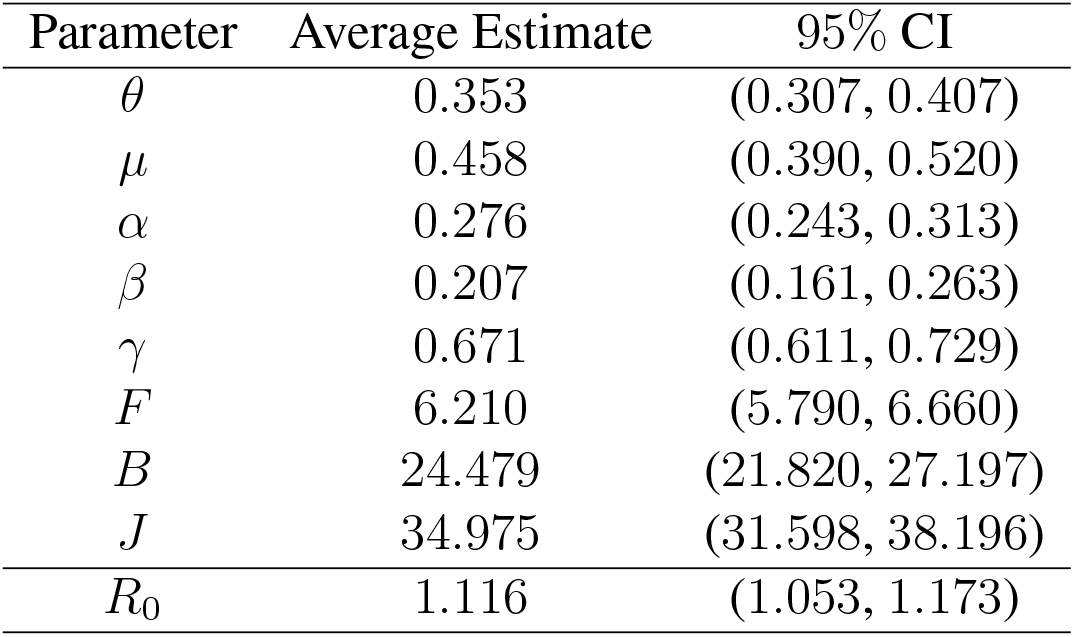
The average estimates and 95% CIs for the parameters

### 4.3 Dynamic Projection of Subpopulation Sizes

To project possible trajectories of the pandemic in Quebec, Canada, we are interested in visualizing the estimated sizes for the six subpopulations in the next 1000 days. To this end, let 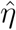 denote the vector of the parameter estimates reported in Table 2. Let 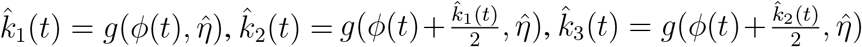, and 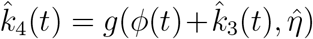. Then similar to (11), we apply the RK4 method to estimate *ϕ*(*t*) recursively for *t* = *τ*_0_ + 1, *τ*_0_ + 2, …, *τ*_0_ + 1000:

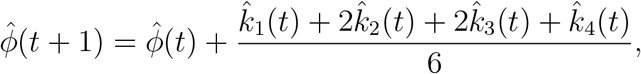

where the initial value *ϕ*(*τ*_0_) is given in Section 3.1.

Figure 3 presents smooth lines connecting the estimates of an element of 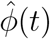 for the time points from *t* = *τ*_0_ to *t* = *τ*_0_+1000, where *S*(*τ*_0_) is roughly equal to 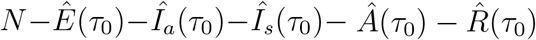, with *Ê*(*τ*_0_), *Îa*(*τ*_0_), *Îs*(*τ*_0_), *Â*(*τ*_0_) and 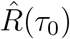 respectively denoting estimates for the corresponding subpopulation size and *N* being the population size of Quebec. Figure 3 shows the patterns that may be useful for us to project the pandemic evolution in Quebec in the next 1000 days if no interference measures are taken. The susceptible subpopulation size *S*(*t*) decreases monotonically, suggesting that as time goes by, more people would move to the subpopulation of exposed cases. *E*(*t*), *I*_*a*_(*t*), *I*_*s*_(*t*) and *A*(*t*) have similar shape with a single peak before 250 days. The size *R*(*t*) of removed cases is an increasing function of time *t*, and it becomes fairly flat after 600 days.

### 4.4 Prediction of Confirmed Cases

With the estimated parameters for the SEASAR model using the data from April 2, 2020 to April 30, 2020, we now predict the daily number of cases for the period of May 1, 2020 to May 10, 2020, and also compare them to the actually reported daily cases in this period. For each *i*, the *i*th estimate and prediction are generated as follows:

- **Step 1:** Let 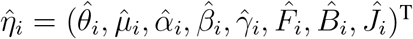 denote the *i*th estimate of parameters, then 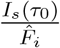 is the *i*th estimate of the number of cases on April 2, 2020.
- **Step 2:** Let 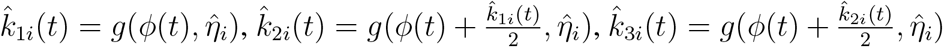 and 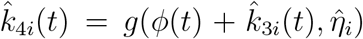, then similar to (11), the *i*th estimate of the sizes of the six subpopulations at the beginning of April 3, 2020, denoted 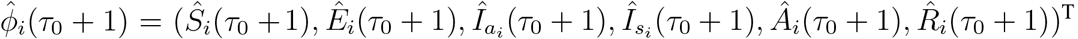, is given by

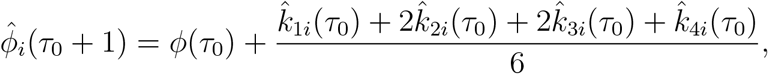

and thus, 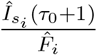 is taken as the *i*th estimate of the number of cases on April 3, 2020.

Repeating Step 2 for *τ*_0_ + 2, …, *τ*_0_ + 28, we obtain the *i*th estimate of the daily number of cases from April 2, 2020 to April 30, 2020. Further repeating Step 2 for *τ*_0_ + 29, …, *τ*_0_ + 38, we obtain the *i*th prediction of the daily number of cases from May 1, 2020 to May 10, 2020.

To evaluate the differences between the predicted and reported daily number of cases from May 1 to May 10, we calculate the *mean absolute error* (MAE), 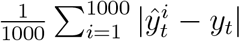, and the *relative mean absolute error* (RMAE), 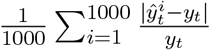, for day *t*, where for *i* = 1, …, 1000, 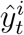 stands for the predicted number of cases on day *t*, and *y*_*t*_ is the reported number of cases on day *t*, defined in Section 3.3. The results are reported in Table 3. Furthermore, Figure 5 displays the mean estimates of the cumulative number of cases for the period of April 2, 2020 to April 30, 2020 (in green) and the mean predicted cumulative number of cases for the period of May 1, 2020 to May 10, 2020 (in blue), in comparison to the actually reported cumulative number of cases from April 2, 2020 to May 10, 2020 (in red). The fair agreement of the fitted or predicted values to the reported values suggest the good performance of the proposed SEASAR model.

**Table 3:**
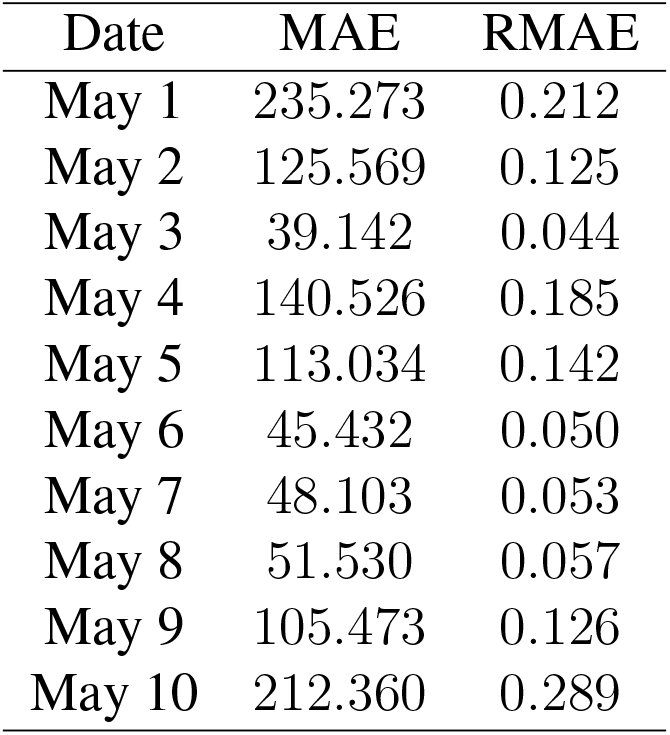
Differences between the predicted and reported numbers

**Figure 5:**
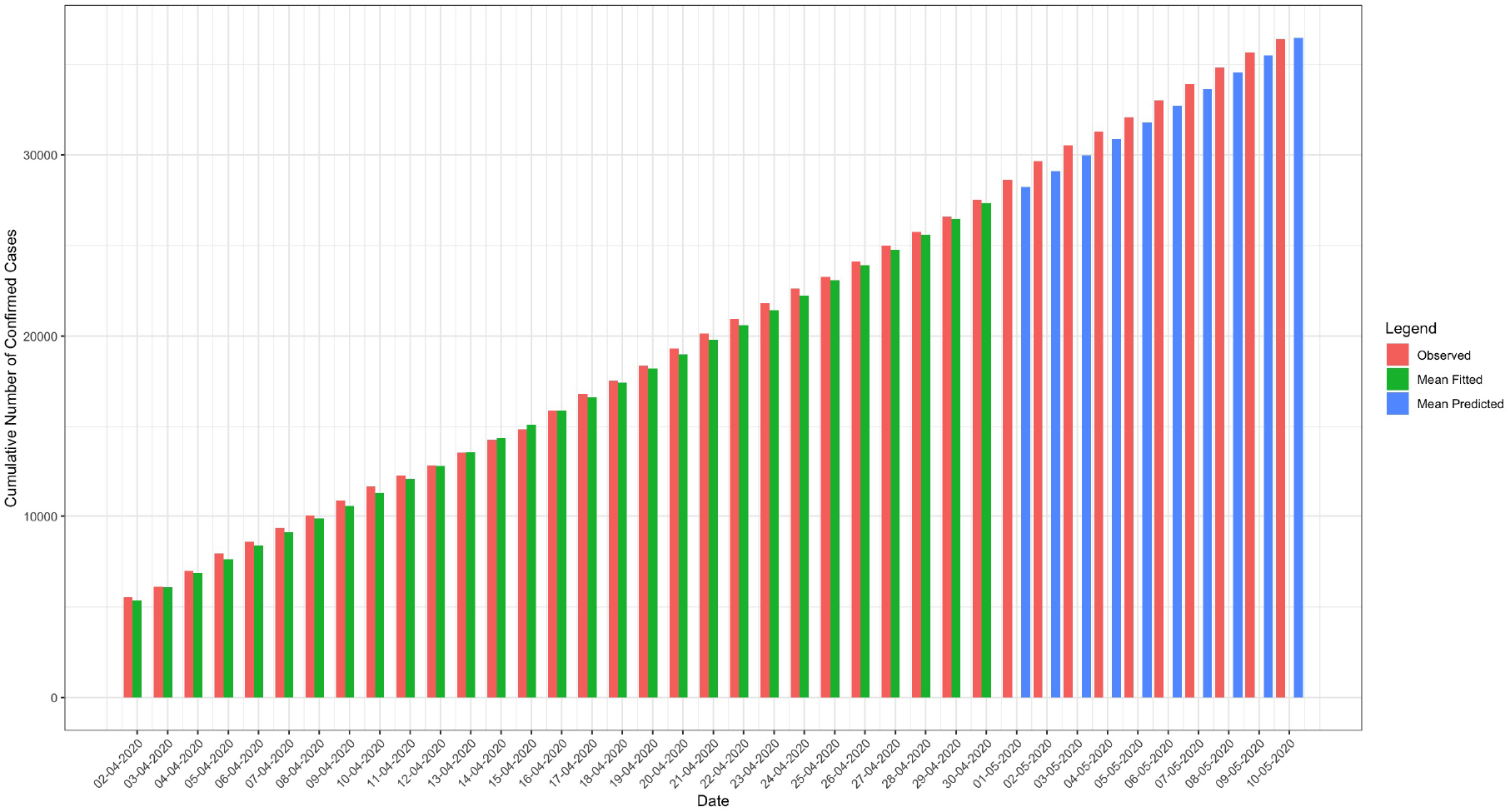
The model fitting to the cumulative number of cases for each day (in green) in the period of April 2, 2020 to April 30, 2020, and the model prediction to the cumulative number of cases for each day (in blue) in the period of May 1, 2020 to May 10, 2020, as opposed to the reported cumulative number of cases for each day (in red) in the period of April 2, 2020 to May 10, 2020.

### 4.5 Sensitivity Analysis

To further assess the performance of the proposed SEASAR model, we conduct ten sensitivity analyses to evaluate the sensitivity of the results to the specification of the initial values. We take the same setup for Section 4.2 except for changing one value as one of the following settings: (**S1**): The observational error variance (OEV) is taken as: 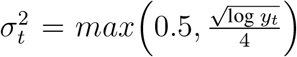; (**S2**): The latent period is set as *Z* = 4.0 days (Guan et al., 2020); (**S3**): The latent period is taken as *Z* = 5.08 days (He et al., 2020); (**S4**): The latent period is set as *Z* = 6.4 days (Backer et al., 2020); (**S5**): Set *r*_3_ = 1; (**S6**): Set *r*_3_ = 3; (**S7**): Set *r*_2_ = 2; (**S8**): Set *r*_2_ = 0.5; (**S9**): Set *r*_1_ = 0.1; (**S10**): Set *r*_1_ = 3.

The results of sensitivity analyses are reported in Figure 6. While the disparity of the fitted values or predicted values from the reported values varies from setting to setting, overall, those differences seem to be fairly small, suggesting reasonable robustness of the results to the specification of associated values in fairly realistic ranges. Further, we report in Table 4 the estimate and associated 95% CIs of the basic reproduction number *R*_0_ obtained from those ten sensitivity analyses. All the estimates are greater than 1, and consequently, the proposed model suggests the ongoing virus spread under a variety of settings we consider.

**Table 4:**
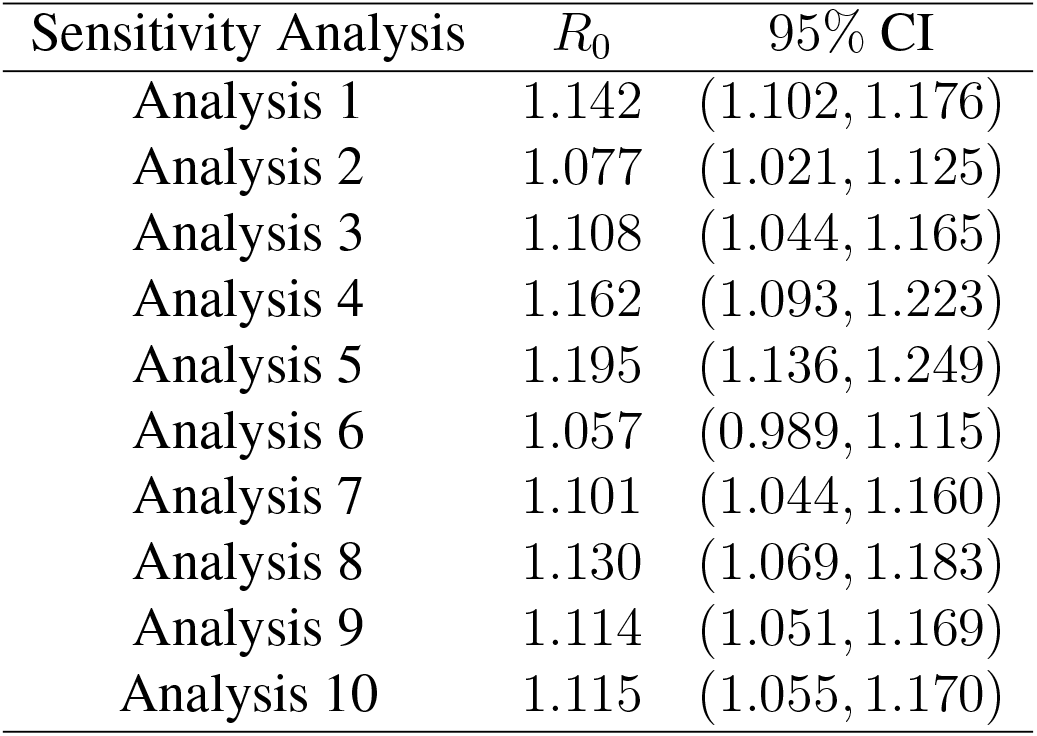
*R*_0_ and the corresponding 95% CIs for 10 sensitivity analyses

| Sensitivity Analysis | $R_0$ | 95% CI |
| --- | --- | --- |
| Analysis 1 | 1.142 | (1.102, 1.176) |
| Analysis 2 | 1.077 | (1.021, 1.125) |
| Analysis 3 | 1.108 | (1.044, 1.165) |
| Analysis 4 | 1.162 | (1.093, 1.223) |
| Analysis 5 | 1.195 | (1.136, 1.249) |
| Analysis 6 | 1.057 | (0.989, 1.115) |
| Analysis 7 | 1.101 | (1.044, 1.160) |
| Analysis 8 | 1.130 | (1.069, 1.183) |
| Analysis 9 | 1.114 | (1.051, 1.169) |
| Analysis 10 | 1.115 | (1.055, 1.170) |

**Figure 6:**
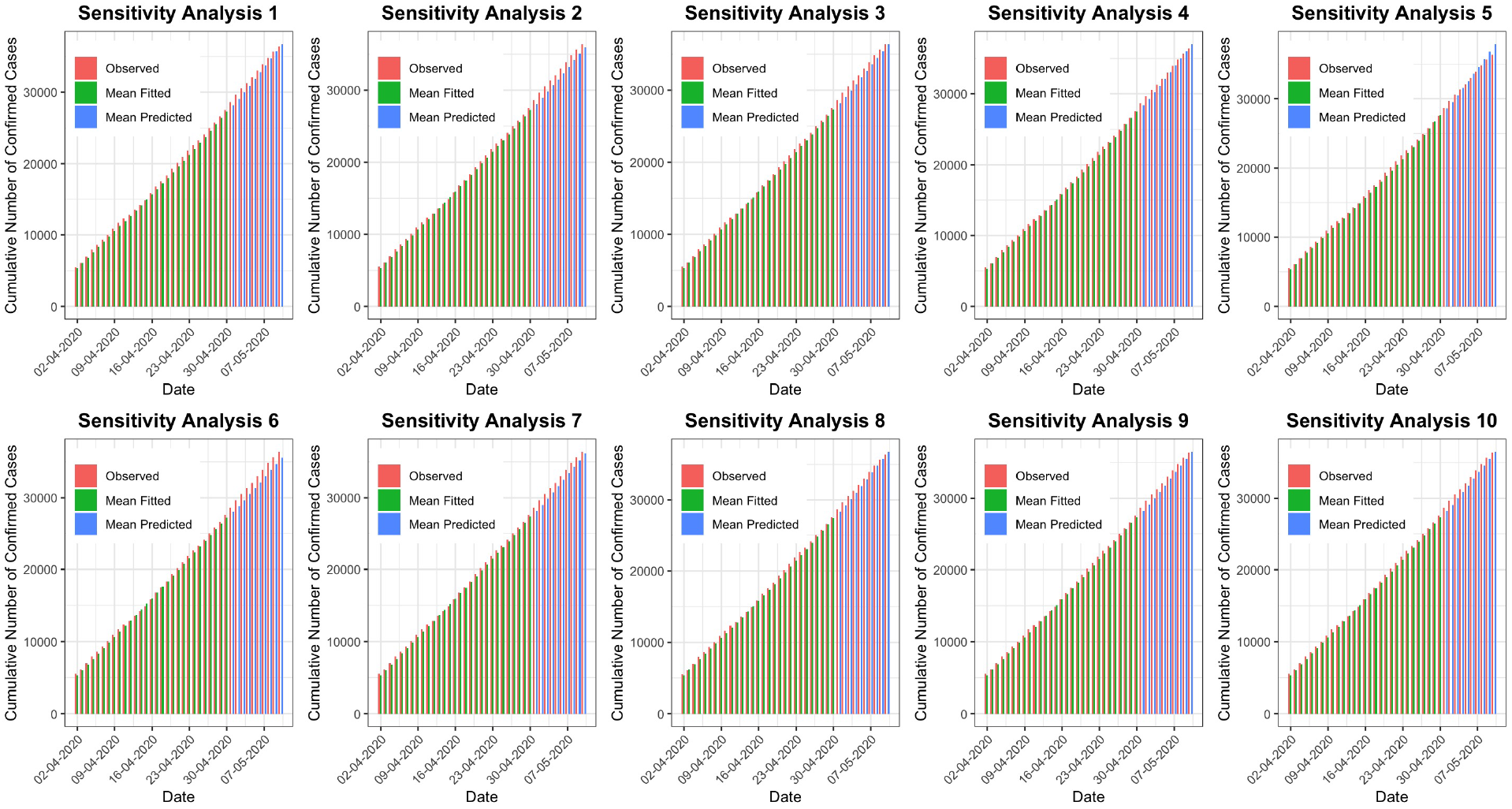
The model fitting to the cumulative number of cases for each day (in green) in the period of April 2, 2020 to April 30, 2020, and the model prediction to the cumulative number of cases for each day (in blue) in the period of May 1, 2020 to May 10, 2020, as opposed to the reported cumulative number of cases for each day (in red) in the period of April 2, 2020 to May 10, 2020.

The results of sensitivity analyses are reported in Figure 6. While the disparity of the fitted values or predicted values from the reported values varies from setting to setting, overall, those differences seem to be fairly small, suggesting reasonable robustness of the results to the specification of associated values in fairly realistic ranges. Further, we report in Table 4 the estimate and associated 95% CIs of the basic reproduction number *R*_0_ obtained from those ten sensitivity analyses. All the estimates are greater than 1, and consequently, the proposed model suggests the ongoing virus spread under a variety of settings we consider.

The results of sensitivity analyses are reported in Figure 6. While the disparity of the fitted values or predicted values from the reported values varies from setting to setting, overall, those differences seem to be fairly small, suggesting reasonable robustness of the results to the specification of associated values in fairly realistic ranges. Further, we report in Table 4 the estimate and associated 95% CIs of the basic reproduction number *R*_0_ obtained from those ten sensitivity analyses. All the estimates are greater than 1, and consequently, the proposed model suggests the ongoing virus spread under a variety of settings we consider.

## 5 Discussion

In this paper, we propose a new epidemic model, called the SEASAR model, to describe the transmission process of COVID-19. Consistent with many available epidemic models, our proposed model require two standard conditions: (1) the population is homogeneous and well-mixed; and (2) the population size remains invariant over time. To facilitate different states of the individuals in the population, we divide the population into six subpopulations (or compartments), respectively, called *susceptible, exposed, asymptomatic, symptomatic, active*, and *removed*. Such a classification allows us to accommodate the unique manifestations of COVID-19 which include asymptomatic infections and varying lag times between symptoms onset and diagnosis. The dynamic changes from compartment to compartment is delineated by ordinary differntial equations together with unknown parameters or transition rates.

To utilize the proposed model, we examine the COVID-19 data in Quebec, Canada, from April 2, 2020 to May 10, 2020. Our analysis is conducted under varying assumptions to reflect different possibilities due to the lack of the precise information. The sensitivity analyses reveal that the pandemic situation in Quebec, Canada, is not under control for the study period.

The proposed SEASAR model extends several existing epidemic models such as SIR and SEIR models. It would be interesting to develop more refined models along the same lines of the current development. For example, one may relax constant model parameters to be time-varying to gain a greater flexibility. Time-varying model parameters may be described by a parametric form or a weakly parametric form (e.g., piecewise constants over different time intervals). While the same principle can be applied to develop a procedure for estimating model parameters, technical details would be more notationally involved, and computation would be more costly.

## Data Availability

The data set used is available publicly.

## Acknowledgements

This research is partially supported by the Natural Sciences and Engineering Research Council of Canada (NSERC) as well as the Rapid Response Program COVID-19 of the Canadian Statistical Sciences Institute (CANSSI). Yi is Canada Research Chair in Data Science (Tier 1). Her research was undertaken, in part, thanks to funding from the Canada Research Chairs Program.

## Appendix A: Expression of the Basic Reproduction Number

Following the principles of Diekmann et al. (2010), we derive the *basic reproduction number R*_0_. The basic idea is to extract the information on the production of new infections from the model, and then calculate the expected number of new infections generated from a case in the population under the constraint *S*(*t*) = *N*.

Since a confirmed case must be quarantined, any individuals in the compartment of being active cannot be infectious. In addition, any individuals in the compartments of being susceptible or removed do not have infectious abilities. Therefore, only (2), (3) and (4) in the SEASAR model involves the information on new infections generated by infected cases who are not confirmed yet, thus not being quarantined. With the assumption that all individuals in the populations are susceptible (i.e., setting *S*(*t*) = *N*), those three equations become

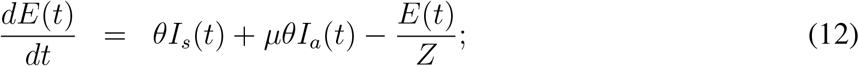

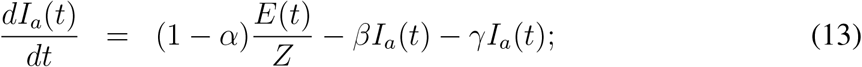

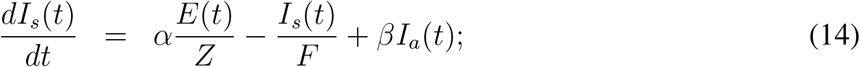

which can be equivalently written in a compact form:

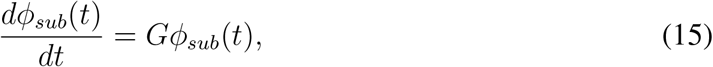

where *ϕ*_*sub*_(*t*) = (*E*(*t*), *I*_*a*_(*t*), *I*_*s*_(*t*))^T^ and 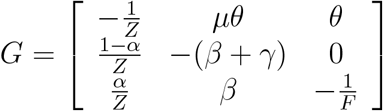.

Noting that only equation (12) includes the terms, *θI*_*s*_(*t*)+*µθI*_*a*_(*t*), of the production of new infections, we re-write the matrix *G* as *G* = *G*_1_ + *G*_2_, where 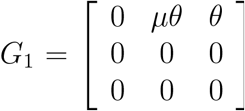 reflects the coefficients in the expression *θI*_*s*_(*t*)+*µθI*_*a*_(*t*), and 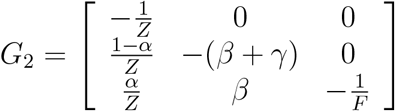 Although the definition of *R*_0_ is conceptually intuitive, its determination is mathematically complicated (Delamater et al., 2019). Diekmann et al. (1990) took *R*_0_ as the dominant eigenvalue of the next-generation matrix, which is derived as 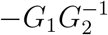 for the equations in (15) by adapting the arguments of Diekmann et al. (2010). Specifically, noting that 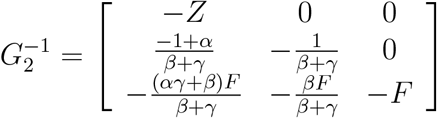, we obtain that 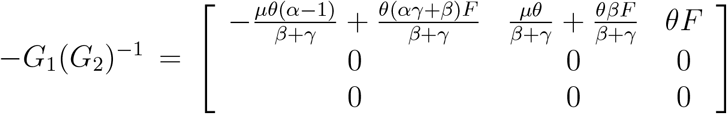, whose dominant eigenvalue is 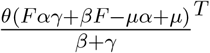. Therefore, 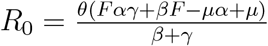.

## Appendix B: Derivations of the Expressions in Section 3.3

In this appendix, we present the derivations of the expressions used in Section 3.3, and describe the implementation of the IF-EAKF algorithm using the terminology of the “joint state-observation vector” (see Anderson, 2001, p.2886) which basically includes time-dependent variables.

For ease of exposition, we let log *ϕ*(*t*) denote the vector (log *S*(*t*), log *E*(*t*), log *I*_*a*_(*t*), log *I*_*s*_(*t*), log *A*(*t*), log *R*(*t*))^T^. Let the joint state-observation vector at time *t* be *O*(*t*) = (*η*^T^, (log *ϕ*(*t*))^T^, log *Ŷ*(*t*))^T^. Because *ϕ*(*t*) is regarded as deterministic at *τ*_0_, the beginning of the study, so when *t* = *τ*_0_, *O*(*t*) degenerates as *O*(*τ*_0_) = (*η*^T^, log *Ŷ*(*τ*_0_))^T^. Here applying the logarithm to *ϕ*(*t*) and *Ŷ*(*t*) aligns with the transformation involved with model (7).

The EAKF algorithm basically aims to work out the posterior distribution of the joint state-observation vector *O*(*t*) for *t* ≥ *τ*_0_. To reduce computation costs, we adopt the discussion of (Anderson, 2001, p.2888) and consider a simple implementation procedure by examining *O*(*t*) via its paired subvectors separately, where we pair log *Ŷ*(*t*) with each of other elements in *O*(*t*) and calculate the posterior distributions for (*θ*, log *Ŷ*(*t*))^T^, (*µ*, log *Ŷ*(*t*))^T^, (*α*, log *Ŷ*(*t*))^T^, (*β*, log *Ŷ*(*t*))^T^, (*γ*, log *Ŷ*(*t*))^T^, (*F*, log *Ŷ*(*t*))^T^, (*B*, log *Ŷ*(*t*))^T^, (*J*, log *Ŷ*(*t*))^T^, (log *S*(*t*), log *Ŷ*(*t*))^T^, (log *E*(*t*), log *Ŷ*(*t*))^T^,(log *I*_*a*_(*t*), log *Ŷ*(*t*))^T^, (log *I*_*s*_(*t*), log *Ŷ*(*t*))^T^, (log *A*(*t*), log *Ŷ*(*t*))^T^, and (log *R*(*t*), log *Ŷ*(*t*))^T^ separately, which employs the same procedure in principle.

To show the ideas, we just describe the way of calculating the posterior distribution of (*θ*, log *Ŷ*(*t*))^T^ for *t* ≥ *τ*_0_. Write *Z*_*t*_ = (*θ*, log *Ŷ*(*t*))^T^, and let *H* = (0, 1) so that *HZ*_*t*_ = log *Ŷ*(*t*). Rather than attempt to find the analytic expression of the posterior distribution, the EAKF algorithm generates the posterior values of *Z*_*t*_ by using the prior values of *Z*_*t*_ and the observation *y*_*t*_. To this end, let 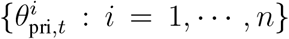 and 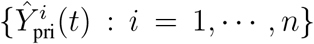 denote sequences of prior values of *θ* and *Ŷ*(*t*) at time *t*, respectively.

First, we calculate their sample means, sample covariance, and sample variances, which are, respectively, given by

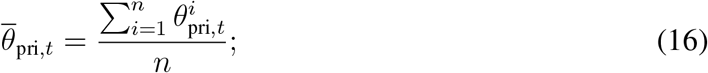

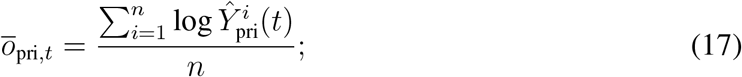

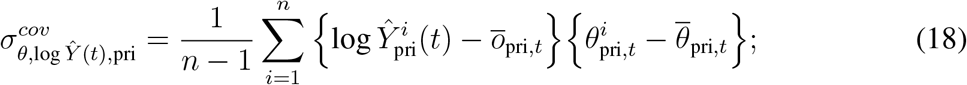

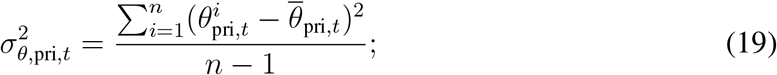

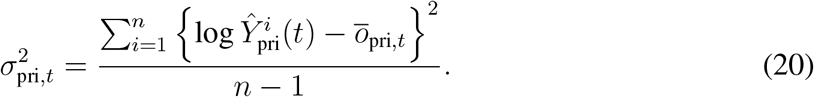

Then we set the mean of prior values of *Z*_*t*_ as

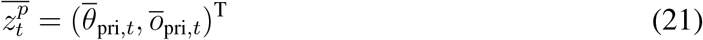

and the covariance matrix of prior values of *Z*_*t*_ as

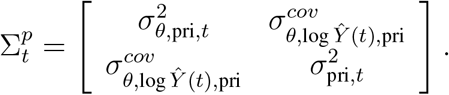

The EAKF algorithm assumes that the prior distribution of *Z*_*t*_ can be represented or reasonably approximated by a Gaussian distribution with the mean 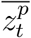 and the covariance matrix 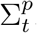. Therefore, the density function of the prior distribution of *Z*_*t*_ is expressed as

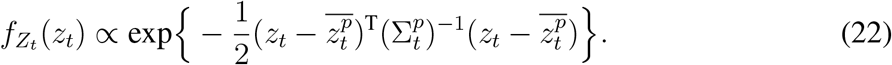

In Appendix C, we show that the posterior distribution of *Z*_*t*_ is Gaussian with the mean

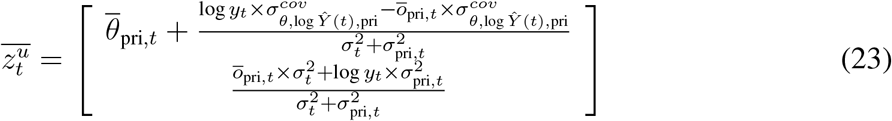

and covariance matrix

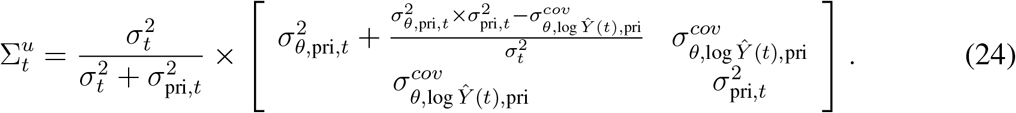

To describe the generation of the posterior values of *θ* and *Ŷ*(*t*) at time *t*, for *i* = 1, …, *n*, we let 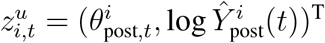 denote posterior values of *θ* and log *Ŷ*(*t*) to be generated, and let 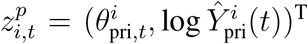 denote prior values of *θ* and log *Ŷ*(*t*). Then the EAKF algorithm (Anderson, 2001, p.2887) generates *n* posterior values of *Z*_*t*_ as follows:

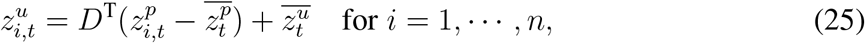

where 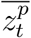 and 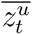 are given by (21) and (23), respectively, and *D* can be any matrix such that the covariance matrix of 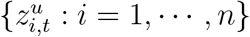 computed from (25) equals that computed by (24).

The existence of such a matrix *D* is justified in Appendix A of Anderson (2001). For instance, *D* can be taken as

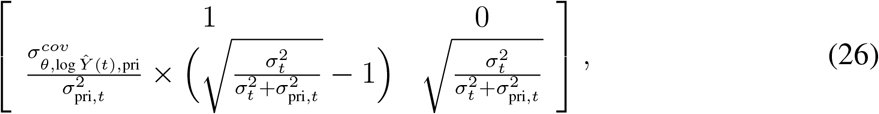

as shown in Appendix D.

Substituting matrix *D* in (25) with (26), we obtain the following explicit expressions for posterior values of *θ* and log *Ŷ*(*t*) at time *t* for *i* = 1, …, *n*:

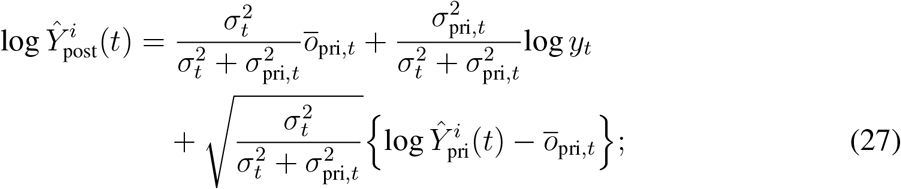

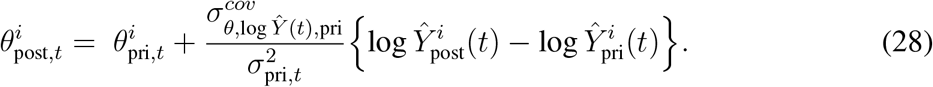

Setting *t* = *τ*_0_, (27) and (28) yield (9) and (10), respectively.

## Appendix C: Derivations of the Posterior Distribution of *Z*_*t*_

For ease of notation, we write *U* = log *Y* (*t*) and *Z* = *Z*_*t*_ for any given *t* ≥ *τ*_0_. Let *H*_1_ = (1, 0), by the fact that *Z* = (*θ*, log *Ŷ*(*t*))^T^ and *H* = (0, 1), we have *θ* = *H*_1_*Z* and log *Ŷ*(*t*) = *HZ*.

Now we show that *U* is conditionally independent of *H*_1_*Z*, given *HZ*. Indeed, for any *u, z*_1_, *z*_2_ ∈ ℝ, the conditional cumulative distribution function of *U* and *H*_1_*Z*, given *HZ* = *z*_2_, is

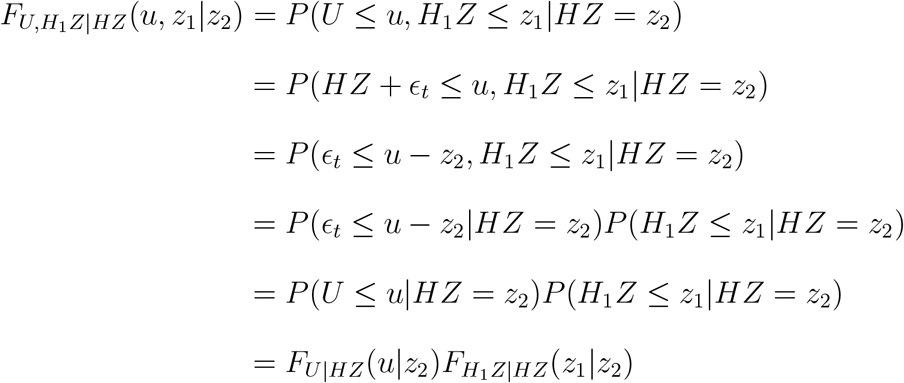

where the second equality is due to (7), the fourth equality comes from the assumption that *ϵ*_*t*_ is independent of *H*_1_*Z*, and the second last step is because that *P* (*ϵ*_*t*_ ≤ *u* − *z*_2_|*HZ* = *z*_2_) = *P* (*ϵ*_*t*_ ≤ *u* − *HZ*|*HZ* = *z*_2_) = *P* (*HZ* + *ϵ*_*t*_ ≤ *u*|*HZ* = *z*_2_) = *P* (*U* ≤ *u*|*HZ* = *z*_2_) by (7). Therefore, we conclude that *U* is independent of *H*_1_*Z*, given *HZ*.

Next, we find the conditional distribution of *U* given *Z, f*_*U*|*Z*_(*u*|*z*), where *z* = (*z*_1_, *z*_2_)^T^ with *z*_1_, *z*_2_ ∈ ℝ, and *u* ∈ ℝ. To do this, we use the definition,

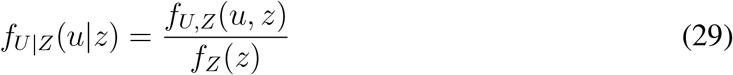

and first determine the joint distribution of *U* and *Z, f*_*U,Z*_(*u, z*). By the definition of *Z*, we have

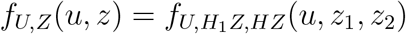

which equals 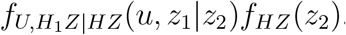. Since *U* is conditionally independent of *H*_1_*Z*, given *HZ*, therefore, we have that

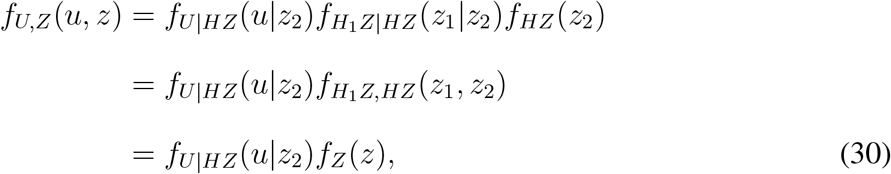

where the last equality is due to the definition of *Z* = (*H*_1_*Z, HZ*)^T^. Combining (30) and (29) gives that

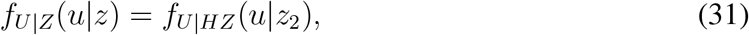

which is determined by (7). Therefore, using the original notation, by (31), we obtain the conditional density function of log *Y* (*t*) given *Z*_*t*_,

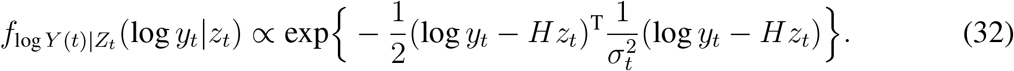

Finally, we determine the posterior distribution of *Z*, i.e., the conditional distribution of *Z* given *U*, *f*_*Z*|*U*_ (*z*|*u*). Combining (22) and (32) gives the density function of the posterior distribution of *Z*_*t*_:

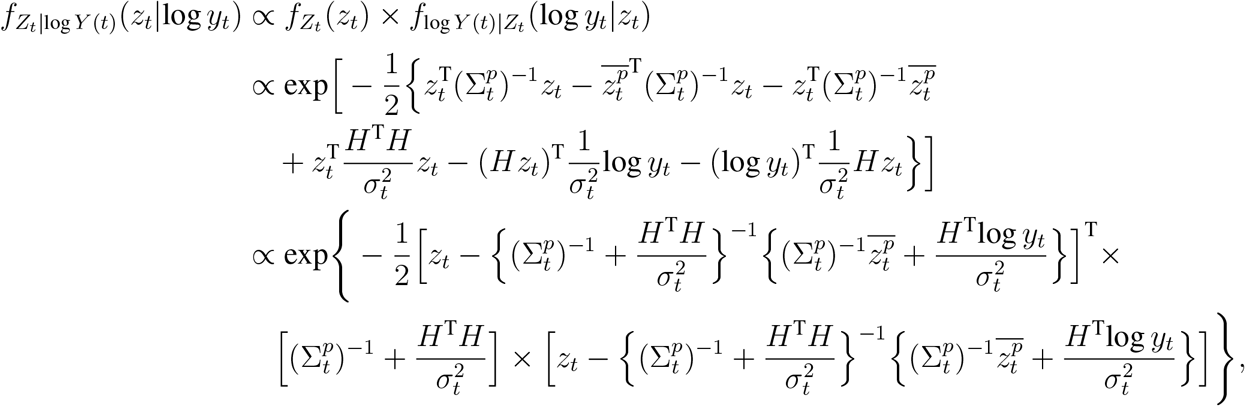

showing that the posterior distribution of *Z*_*t*_ is Gaussian with the mean

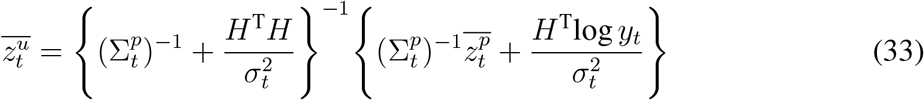

and covariance matrix

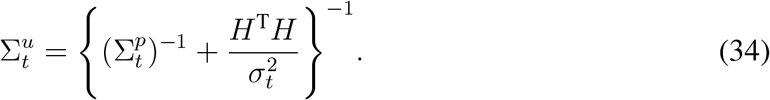

Furthermore, we express (33) and (34) in terms of the observations and the prior values in (16), (17), (18), (19), and (20):

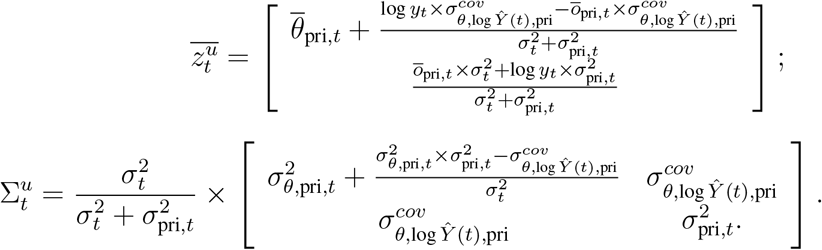

## Appendix D: Verification of the Condition Required from EAKF

Substituting the matrix *D* in (25) with (26) gives us

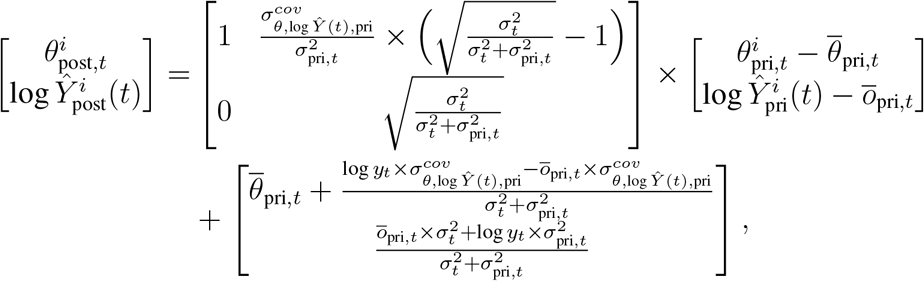

yielding that

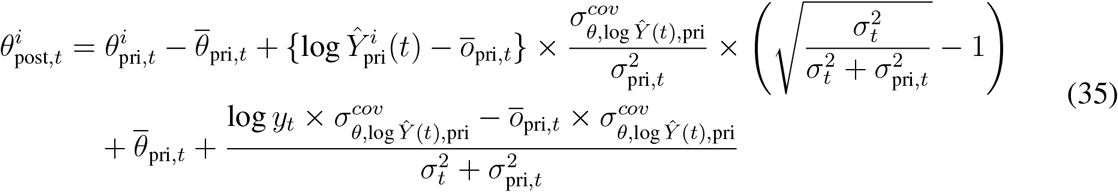

and

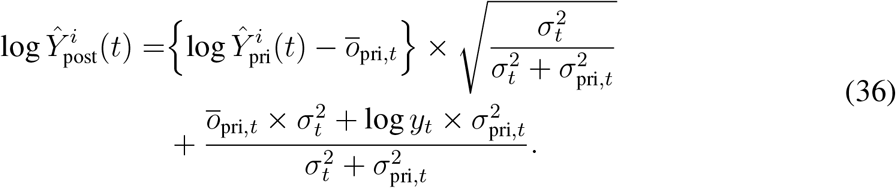

Thus, (35) and (36) give us a sequence of posterior values of 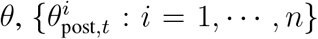, and a sequence of posterior values of 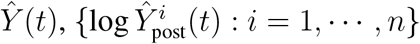.

Next, we calculate the variance of the posterior values 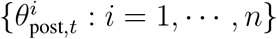 using (35),

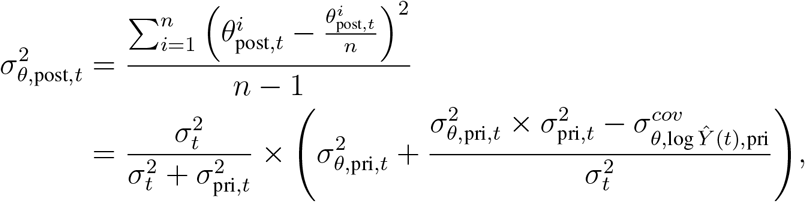

which is identical to the (1, 1) element of (24).

Using (36), we calculate the variance of the posterior values 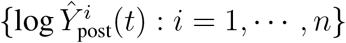 of log *Ŷ*(*t*), given by

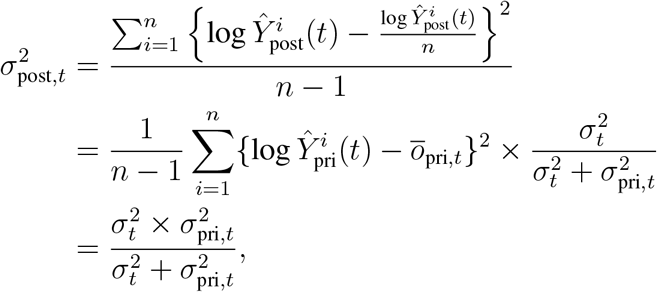

which is identical to the (2, 2) element of (24).

Furthermore, using (35) and (36), we calculate the covariance of 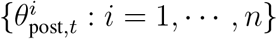 and 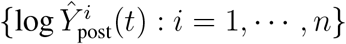,

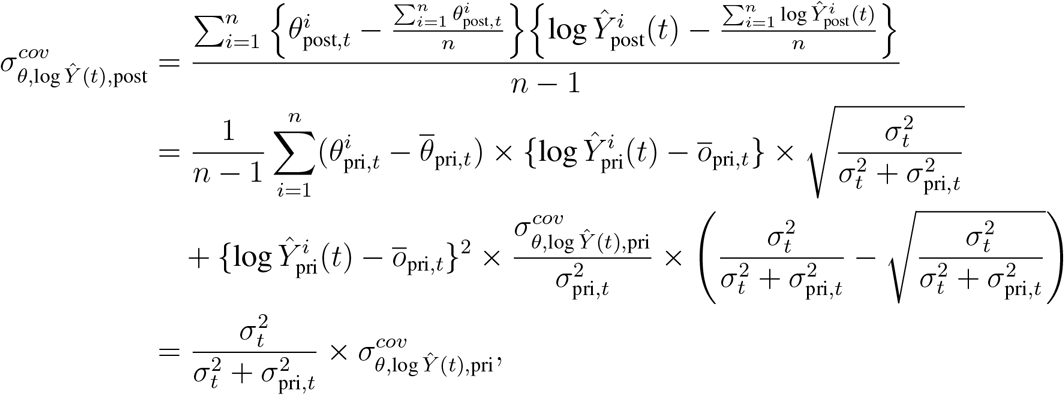

which is identical to the (1, 2) and (2, 1) elements of the matrix (24). Therefore, we verify that the matrix *D* satisfies the condition required by the EAKF algorithm.

